# Unravelling the link between sleep and mental health during the COVID-19 pandemic

**DOI:** 10.1101/2022.03.28.22273027

**Authors:** Juan González-Hijón, Anna K. Kähler, Emma M. Frans, Unnur A. Valdimarsdóttir, Patrick F. Sullivan, Fang Fang, Anikó Lovik

**Affiliations:** Institute of Environmental Medicine, Karolinska Institutet, Solna, Sweden; Department of Medical Epidemiology and Biostatistics, Karolinska Institutet, Solna, Sweden; Center of Public Health Sciences, University of Iceland, Reykjavik, Iceland; Department of Epidemiology, Harvard T. H. Chan School of Public Health, Boston, MA, USA; Department of Psychiatry, University of North Carolina, Chapel Hill, NC, USA; Department of Genetics, University of North Carolina, Chapel Hill, NC, USA

**Keywords:** Anxiety, COVID-19, Depression, Mental Health, Omtanke2020 study, Sleep, Stress, Sweden

## Abstract

**Background:** The emergence of COVID-19 brought unparalleled changes in people’s lifestyle, including sleep. We aimed to assess the bidirectional association between sleep quality and mental health and describe how sleep and mental health were affected in Sweden during the COVID-19 pandemic (between June 2020 and September 2021).

**Methods:** Data were obtained from the Omtanke2020 study. Participants who completed the baseline survey and 8 monthly follow-up surveys were included (N=9035). We described the distribution of sleep and mental health in the different Swedish regions using maps and over the study period with longitudinal graphs adjusting for sex, age, recruitment type (self-recruitment or invitation), and COVID-19 status. The inner relationships between mental health, sleep and covid infection were described through relative importance networks. Finally, we modelled how mental health affects sleep and vice versa using generalized estimating equations with different adjustments.

**Results:** Seasonal and north-south regional variations were found in sleep and mental health outcomes at baseline and attenuated over time. The seasonal variation of sleep and mental health correlated moderately with the incidence rate of COVID-19 in the sample. Networks indicate that the relationship between COVID-19 incidence and mental health varies over time. We observed a bidirectional relationship between sleep quality and quantity at baseline and mental health at follow-up and vice versa.

**Conclusion:** Sleep quality and quantity at baseline was associated with adverse symptom trajectories of mental health at follow-up, and vice versa, during the COVID-19 pandemic. There is also a weak relationship between COVID-19 incidence, sleep, and mental health.

## Introduction

The outbreak of the SARS-CoV-2 virus infection at the end of 2019 is one of the most wide-spread life-changing events globally in recent history. On the one hand, people have lived with the preoccupation of being infected, themselves or their loved ones, with a virus that, for some, proves to be a potentially deadly disease (Boyraz, Legros, & Tigershtrom, 2020). On the other hand, to stop the spread of the virus, most governments have adopted Non-Pharmaceutical Intervention (NPI) measures (Ferguson et al., 2020), such as lockdowns, working from home policies, mandatory facemask wearing, etc. During the pandemic, many have experienced great changes in communication, work, or even leisure activities due to these measures.

Exposure to mass disasters is related to an increase in the risk of adverse mental health symptoms, including post-traumatic stress disorder (PTSD) (Galea, Nandi, & Vlahov, 2005), depression (Chen et al., 2006), anxiety (Chen et al., 2006), and sleep disorders (Porcheret, Stensland, Wentzel-Larsen, & Dyb, 2022). These mental health problems have been seen in natural disasters such as volcanic eruption (Carlsen et al., 2012), tsunami (Arnberg et al., 2015), and epidemic outbreak (Chen et al., 2006) as well as in human-made crises such as a national economic collapse (Hauksdóttir, McClure, Jonsson, Ólafsson, & Valdimarsdóttir, 2013) and terrorism (Porcheret, Stensland, Wentzel-Larsen, & Dyb, 2022).

Even though Sweden, unlike most countries, was not exposed to a strict lockdown, other measures were indeed taken to stop the spread of COVID-19, including closure of the border, travel restrictions, working from home, remote-based classes in universities and upper secondary schools, participant limits and cancellation of events, closure of public spaces, opening hours and capacity restrictions in hospitals, quarantines, and social distancing (“Lag (2021:4) om särskilda begränsningar för att förhindra spridning av sjukdomen covid-19 Svensk författningssamling 2021:2021:4 - Riksdagen,” 2021). Although these measures may have been effective in stopping the spread, they come with a social cost, such as an increase in loneliness (Okruszek, Aniszewska-Stańczuk, Piejka, Wiśniewska, & Żurek, 2020) and concerns about loss of income and uncertainty about job security (Morin, Carrier, Bastien, & Godbout, 2020). There has indeed been an increase in unemployment during the pandemic, noted as the highest unemployment rate of the last two decades (“Arbetslöshet i sverige,” 2021).

The pandemic has introduced a combination of factors that trigger stress, such as fear of contagion (Liu et al., 2022), change in daily routines (Khawar et al., 2021), loneliness (Okruszek, Aniszewska-Stańczuk, Piejka, Wiśniewska, & Żurek, 2020), and uncertainty about the future (Morin, Carrier, Bastien, & Godbout, 2020). One common cause of sleep problems is the activation of the hypothalamus-pituitary-adrenal (HPA) axis by a stressor (Van Reeth et al., 2000). This activation directly affects the circadian cycle and consequently sleep regulation. Prolonged activation of the HPA axis can lead to anxiety and major depression (Van Reeth et al., 2000). Bi-directional association between depression, anxiety, and sleep disturbances has also been documented (Kalmbach et al., 2018), even after the stressful event has passed (Morin et al., 2009).

In general, the COVID-19 pandemic has led to an increase in stress, anxiety, depression, and sleep disturbances in many regions of the world (Halsøy, Johnson, Hoffart, & Ebrahimi, 2021; Zhang et al., 2022). The purpose of our study is to describe the regional variation and the development of the longitudinal relationships between sleep and mental health as well as between sleep, mental health, and the incidence of COVID-19 in Sweden over a 14-month period of the pandemic using a set of different analyses. First, to study the spatial distribution (Figure 1) the percentage of total score for each questionnaire in each region was calculated and depicted on maps at baseline and after 8 months. Then, the temporal distribution was studied through a series of timeline allowing for comparisons between the different outcomes (Figure 2). In the next step, relative importance networks (RIN) were used to graphically evaluate the relationships between different variables (Figure 3). And finally, to reveal the interaction between sleep and mental health during this period, generalized estimating equations (GEE) models were fitted in both directions (Tables 3 and 4). All these analyses contribute towards getting a deep understanding of the longitudinal changes and the bidirectional link between sleep and mental health during the COVID-19 pandemic focusing on the second and third waves of the pandemic when interventions to stop the spread were stricter.

**Figure 1.**
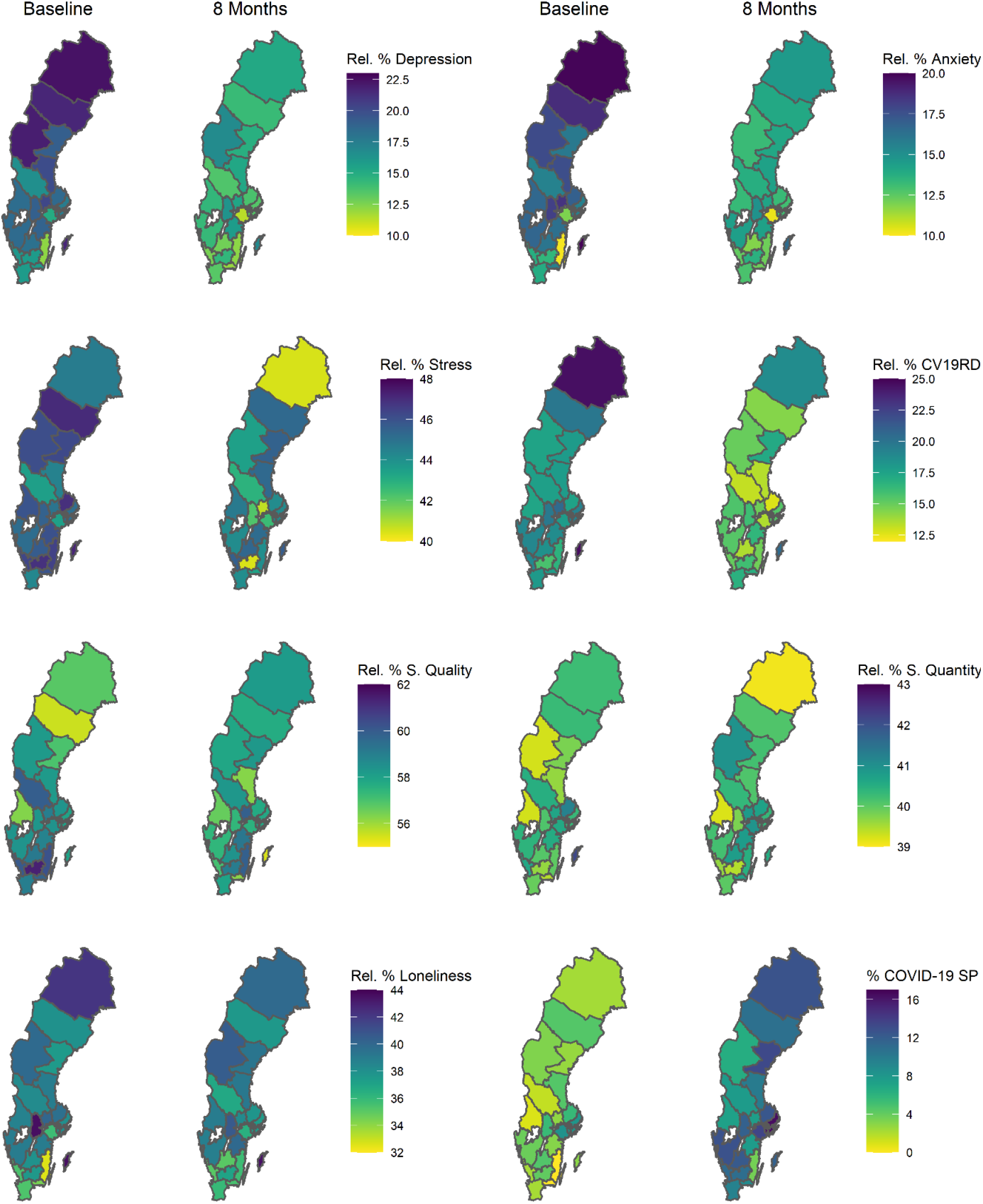
Mean percentages of depression, anxiety, COVID-19-related distress (CV19RD), stress, sleep quality, sleep quantity, loneliness, and COVID-19 incidence, adjusted for sex, age, and recruitment type.

**Figure 2.**
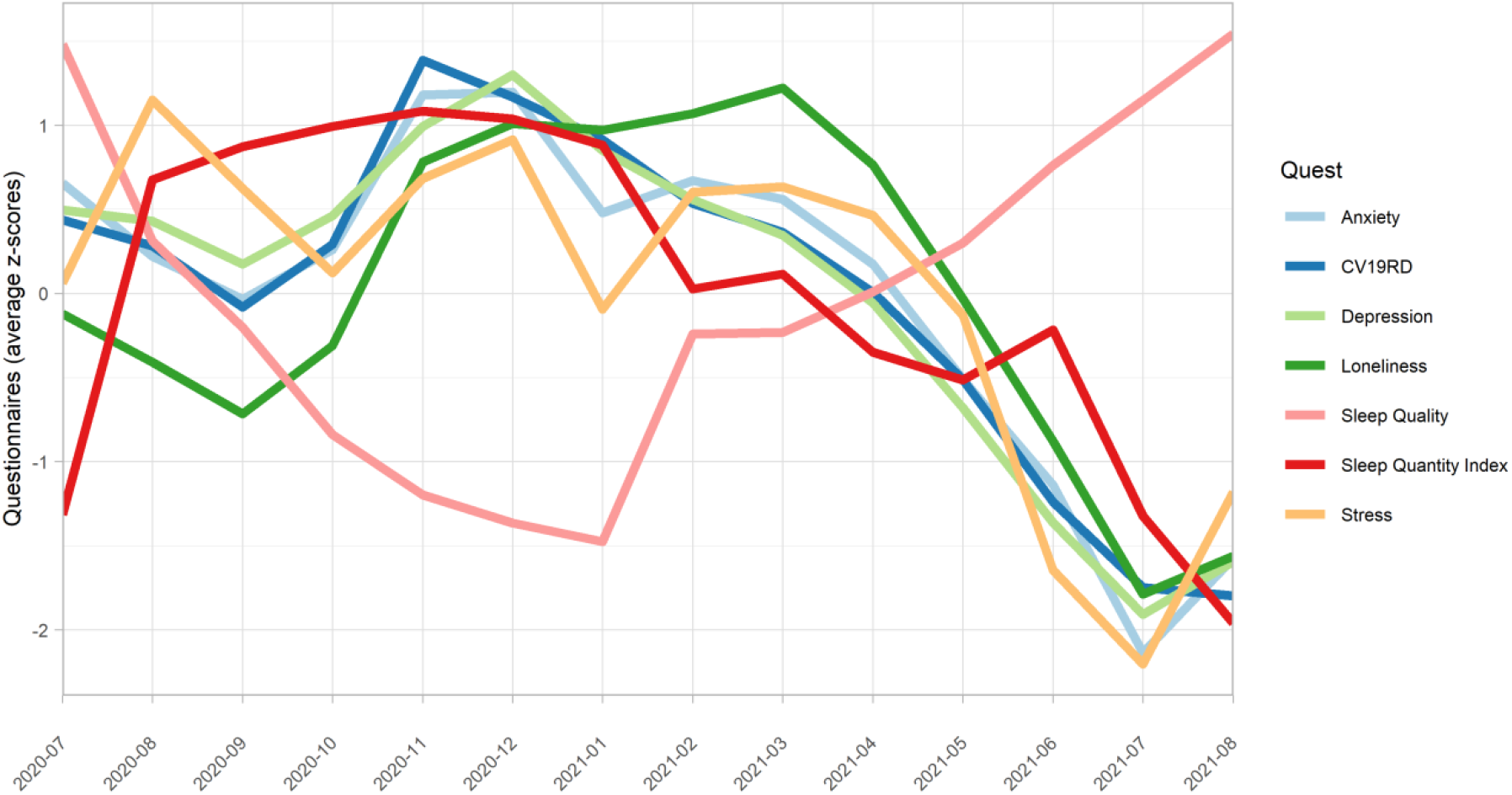
Temporal pattern (average Z-Score) of depression, anxiety, COVID-19-related distress (CV19RD), stress, loneliness, sleep quality, and sleep quantity index, adjusted for age, sex, recruitment type and COVID-19.

**Figure 3.**
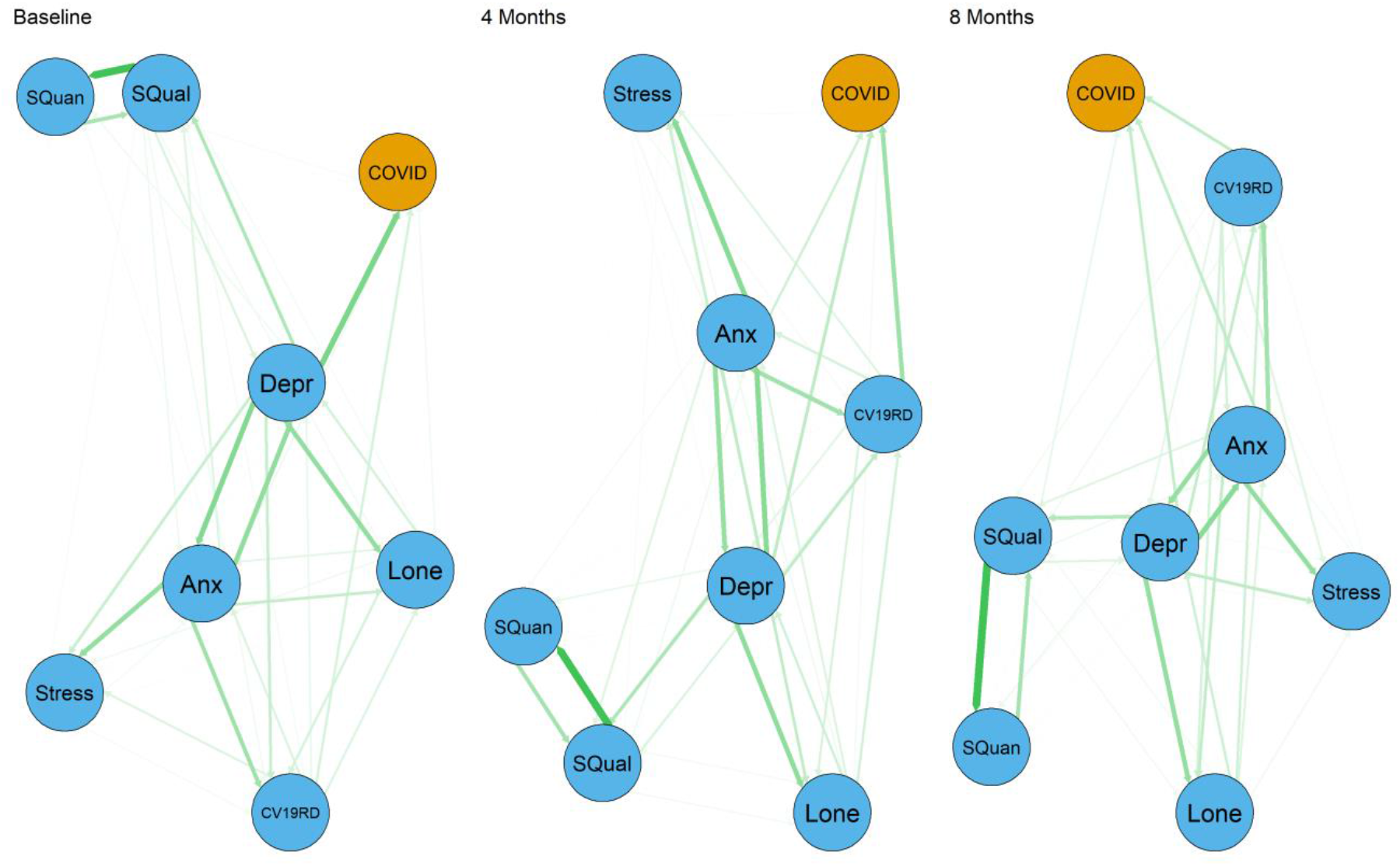
Relationship between depression (Depr), anxiety (Anx), COVID-19-related distress (CV19RD), stress, loneliness (Lone), sleep quality (SQual) and sleep quantity (SQuan) and COVID-19 (COVID).

## Material and Methods

### Participants

In this study, we included in the analysis 9,035 participants who completed the baseline survey as well as at least eight monthly follow-up surveys in the Omtanke2020 study, which is a longitudinal study specifically designed to understand the mental health impact of COVID-19 in Sweden including 27,950 participants (Lovik et al., 2021). All Swedish adults over 18 years of age with a BankID (Swedish digital identity identifier) were eligible to participate in the Omtanke2020 study. Recruitment was carried out through social and traditional media campaigns and invitations to participants of ongoing studies at the Karolinska Institutet. In this study, we only included participants who completed at least 25% of the questions concerning depression, anxiety, Covid-19-related distress, stress, sleep quality, and sleep quantity in the baseline and follow-up surveys.

### Sociodemographic variables

Questions about sex, age, relationships, lifestyle, and general health were asked at baseline, including Body Mass Index (BMI), smoking, excessive alcohol consumption (habitual drinking), previous psychiatric disorders, comorbidities, and having a family member or close friend with COVID-19 (as reported by the participant). Furthermore, participants were asked an additional question about how they were invited to participate in the survey (recruitment type), whether through media (self-recruitment) or by invitation.

### Mental health variables

*Depressive* symptoms were measured using the Patient Health Questionnaire (PHQ-9), a widely used screening tool consisting of nine validated items to assess the presence of depressive symptoms and their severity, measured on a 4-point Likert scale, ranging from 0 (not at all) to 3 (nearly every day). The total score, employed in most analyses, ranges from 0 to 27. A binary variable (yes or no) was defined using a cut-off of 10 for use in Table 3. A PHQ-9 score equal to or greater than 10 has a sensitivity of 88% and a specificity of 88% for major depression (Kroenke, Spitzer, & Williams, 2001).

Symptoms of anxiety were measured by the Generalized Anxiety Disorder scale (GAD-7). This tool consists of seven validated items to measure symptoms of Generalized Anxiety Disorder on a 4-point Likert scale. The total score, used in most analyses, ranges from 0 to 21. A binary variable (yes or no) was defined using a cut-off of 10 for use in Table 3. A GAD-7 score equal or greater than 10 has a sensitivity of 82% and a specificity of 89% for generalized anxiety disorder (Spitzer, Kroenke, Williams, & Löwe, 2006).

*COVID-19-related distress* was measured by an adaptation of the Primary Care PTSD Screen adjusted for COVID-19. This instrument consisted of five items, measured on a 5-point Likert-scale ranging from 0 to 4 and includes three items directly related to COVID-19: “Had nightmares about covid-19?”, “Had thoughts about covid-19 even though you did not want it?” and “Avoided thinking about covid-19 or avoided situations reminiscent of covid-19?”. The total score was used in this study, ranging from 0 to 20. In Table 3, using COVID-19 related distress as a predictor for sleep, a binary variable was created (yes or no) by dichotomizing all items and then applying a cut-off of four, which was shown in an unmodified version with binary items to have 82% specificity and 91% sensitivity (Prins, Bovin, Smolenski, Marx, Kimmerling, et al., 2016).

*Sleep quality* was assessed with a question taken from PSQI (Buysse, Reynolds III, Monk, Berman, & Kupfer, 1989) “In the last 2 weeks, how would you describe your average sleep quality?” Answer to this question was made using a Likert scale from 1 to 5, with the minimum being “Very good” and the maximum “Very poor.” For visualization reasons, in this study, the scale was inverted, with 1 standing for low sleep quality whereas 5 for high sleep quality.

*Sleep quantity* was assessed with a question taken from PSQI (Buysse, Reynolds III, Monk, Berman, & Kupfer, 1989) “In the last 2 weeks, how many hours of sleep have you had in general during one night?” To answer this question, the participant had to enter the number of hours ranging from 0 to 24. Extreme values were discarded, so that we selected a range between 2 to 17 hours. In some analyses, this variable was transformed to the Sleep Quantity Index, an ordinal variable with a range from 0 to 3: 7-9 hours was given a score of “0”, 6 or 10 hours a score of “1”, 5 or 11 hours a score of “2”, and values smaller than 5 or greater than 11 a score of “3.” We used this index to measure the undirected deviance from the most common sleep quantity because the relationship between sleep quantity and mental health is U-shaped (both too little and too much sleep is associated with increased mental health symptoms in similar ways – a sensitivity analysis showing short and long sleep quantity separately can be found in supplementary materials to support this (Tables S4 and S5)).

#### Other mental health variables

Stress was measured by the 4-item version of the Perceived Stress Scale (PSS-4) (Cohen, 1988) whereas loneliness was measured with a single question answered on 5-point Likert scale (Hughes, Waite, Hawkley, & Cacioppo, 2004).

### COVID-19 diagnosis

A COVID-19 diagnosis confirmed by PCR and/or antigen test(s) self-reported by the participants was used to define the status of COVID-19 infection.

### Statistical analysis

#### Missing data

The participants had always the option of not answering a specific question. A score was imputed for the unanswered questions using the MICE package of the R software (Van Buuren & Groothuis-Oudshoorn, 2011), if at least 20% of the questionnaire was answered. The imputation was carried out using the predictive mean matching method, generating a minimum of 32 datasets in each iteration with a maximum number of 50 iterations. After this process, the score for the question was chosen using the mode of these datasets.

#### Descriptive Analysis

Descriptive statistics were adjusted for sex, age, recruitment type, and COVID-19 status.

#### Relative Importance Networks

The analysis was performed using the package “bootnet” (Epskamp, Borsboom, & Fried, 2018), which calculates the RIN by the decomposition of the Pearson’s correlation coefficient (R2) between variables. We used the average method on the orders proposed by Lindeman, Merenda and Gold (lmg) (Groemping, 2006), and, to increase the robustness of the results, the data was bootstrapped 1000 times.

#### Models

To assess the evolution of the mental health and sleep outcomes over time, depending on the sociodemographic and other (mental health or sleep) predictors, generalized estimating equations (GEE) were applied using a Poisson link function. In this analysis, the GEEpack package from R (Højsgaard, Halekoh, & Yan, 2006) was used. Models were fitted for both directions to understand the role of baseline mental health measures to the longitudinal trajectories of sleep quality and quantity as well as the baseline sleep quality and quantity on the longitudinal trajectories of mental health measures. In the former, two types of models were fitted, including one adjusted for sex, age, time, recruitment type, and COVID-19 status and another one additionally adjusted for sociodemographic factors. In the latter, we also adjusted for sex, age, time, recruitment type, and COVID-19 status and additionally for sociodemographic factors.

## Results

Table 1 shows the descriptive of sociodemographic factors among individuals with COVID-19 infection before the study (N=504), individuals who were infected during the study (N=3379), and individuals who were not infected (N=5152). The mean age of the participants was 54.4 years (SD: 15.5 years), with a range from 18 to 94 years. Most of the participants were women (84.3%) but sensitivity analyses stratified by sex show no relevant sex differences in the results (see supplement tables S6 and S7). 57% of the participants were recruited by invitation. Self-recruitment was more common among participants with a COVID-19 infection, compared with those who were not infected. In general, participants with COVID-19 (before or during the study period) were younger and more likely to be in a relationship, to have a higher BMI, to be smoker or habitual drinker, to have previous psychiatric disorders, and to have a relative or friend with COVID-19, compared to participants not infected with COVID-19. On the other hand, people without COVID-19 tended to have a higher number of comorbidities.

**Table 1.** Characteristics of the study participants according to their COVID-19 status.

|  | Not infected with<br>COVID-19 | Infected with COVID-19<br>during the study | Infected with COVID-19<br>before the study | Overall |
| --- | --- | --- | --- | --- |
| **No.** | 5152 | 3379 | 504 | 9035 |
| **Sex** |  |  |  |  |
| Female | 4341 (84.3%) | 2870 (84.9%) | 408 (81.0%) | 7619 (84.3%) |
| Male | 811 (15.7%) | 509 (15.1%) | 96.0 (19.0%) | 1416 (15.7%) |
| **Age, years** |  |  |  |  |
| Mean (SD) | 58.1 (15.2) | 49.4 (14.6) | 50.0 (13.1) | 54.4 (15.5) |
| Median (IQR) | 62.0 (21.0) | 50.0 (22.0) | 51.0 (17.0) | 56.0 (23.0) |
| **Age group, years** |  |  |  |  |
| 18-29 | 342 (6.6%) | 358 (10.6%) | 47.0 (9.3%) | 747 (8.3%) |
| 30-39 | 397 (7.7%) | 590 (17.5%) | 68.0 (13.5%) | 1055 (11.7%) |
| 40-49 | 595 (11.5%) | 670 (19.8%) | 106 (21.0%) | 1371 (15.2%) |
| 50-59 | 1043 (20.2%) | 868 (25.7%) | 169 (33.5%) | 2080 (23.0%) |
| 60-69 | 1385 (26.9%) | 588 (17.4%) | 85.0 (16.9%) | 2058 (22.8%) |
| 70+ | 1390 (27.0%) | 305 (9.0%) | 29.0 (5.8%) | 1724 (19.1%) |
| **Relationship** |  |  |  |  |
| In a relationship | 3660 (71.0%) | 2452 (72.6%) | 360 (71.4%) | 6472 (71.6%) |
| Single | 1474 (28.6%) | 903 (26.7%) | 142 (28.2%) | 2519 (27.9%) |
| Missing | 18.0 (0.3%) | 24.0 (0.7%) | 2.00 (0.4%) | 44.0 (0.5%) |
| **Body Mass Index** |  |  |  |  |
| < 25, Normal & low weight | 2794 (54.2%) | 1797 (53.2%) | 261 (51.8%) | 4852 (53.7%) |
| 25 - 30, Overweight | 1566 (30.4%) | 1020 (30.2%) | 152 (30.2%) | 2738 (30.3%) |
| > 30, Obese | 584 (11.3%) | 425 (12.6%) | 75.0 (14.9%) | 1084 (12.0%) |
| Missing | 208 (4.0%) | 137 (4.1%) | 16.0 (3.2%) | 361 (4.0%) |
| **Current smoking** |  |  |  |  |
| No | 4599 (89.3%) | 2906 (86.0%) | 446 (88.5%) | 7951 (88.0%) |
| Yes | 541 (10.5%) | 467 (13.8%) | 56.0 (11.1%) | 1064 (11.8%) |
| Missing | 12.0 (0.2%) | 6.00 (0.2%) | 2.00 (0.4%) | 20.0 (0.2%) |
| **Habitual drinking\*** |  |  |  |  |
| No | 2798 (54.3%) | 1899 (56.2%) | 287 (56.9%) | 4984 (55.2%) |
| Yes | 1327 (25.8%) | 893 (26.4%) | 137 (27.2%) | 2357 (26.1%) |
| Missing | 1027 (19.9%) | 587 (17.4%) | 80.0 (15.9%) | 1694 (18.7%) |
| **Previous psychiatric disorder** |  |  |  |  |
| No | 3792 (73.6%) | 2213 (65.5%) | 364 (72.2%) | 6369 (70.5%) |
| Yes | 1304 (25.3%) | 1141 (33.8%) | 135 (26.8%) | 2580 (28.6%) |
| Missing | 56.0 (1.1%) | 25.0 (0.7%) | 5.00 (1.0%) | 86.0 (1.0%) |
| **Comorbidities** |  |  |  |  |
| No comorbidity | 3168 (61.5%) | 2386 (70.6%) | 357 (70.8%) | 5911 (65.4%) |
| One comorbidity | 1405 (27.3%) | 759 (22.5%) | 115 (22.8%) | 2279 (25.2%) |
| 2 or more comorbidities | 575 (11.2%) | 233 (6.9%) | 32.0 (6.3%) | 840 (9.3%) |
| Missing | 4.00 (0.1%) | 1.00 (0.0%) | 0 (0%) | 5.00 (0.1%) |
| **Relatives with COVID-19** |  |  |  |  |
| No | 4283 (83.1%) | 2669 (79.0%) | 247 (49.0%) | 7199 (79.7%) |
| Yes | 848 (16.5%) | 694 (20.5%) | 253 (50.2%) | 1795 (19.9%) |
| Missing | 21.0 (0.4%) | 16.0 (0.5%) | 4.00 (0.8%) | 41.0 (0.5%) |
| **Recruitment type (Self-recruited)** |  |  |  |  |
| No | 3086 (59.9%) | 1799 (53.2%) | 262 (52.0%) | 5147 (57.0%) |
| Yes | 1293 (25.1%) | 1106 (32.7%) | 170 (33.7%) | 2569 (28.4%) |
| Missing | 773 (15.0%) | 474 (14.0%) | 72.0 (14.3%) | 1319 (14.6%) |
| **Date of baseline** |  |  |  |  |
| Jun - Aug 2020 | 1713 (33.2%) | 964 (28.5%) | 154 (30.6%) | 2831 (31.3%) |
| Sep - Nov 2020 | 2455 (47.7%) | 1333 (39.4%) | 165 (32.7%) | 3953 (43.8%) |
| Dec 2020 - Feb 2021 | 984 (19.1%) | 1082 (32.0%) | 185 (36.7%) | 2251 (24.9%) |

\*For women, 4 or more drinks consumed on one occasion (one occasion = 2-3 hours). For men, 5 or more drinks consumed on one occasion.

### Mental health and sleep at baseline and eight months

Figure 1 shows the mean percentages of total score in the mental health and sleep measures per each of the 21 regions in Sweden, e.g., with 0% corresponds to the minimum (e.g., 0 for PHQ-9) and 100% to the maximum (e.g., 27 for PHQ-9) of the total score, and percentage of participants with COVID-19. The figure shows percentages at two time points (baseline and at eight monthly follow-up survey) after adjusted for sex, age, recruitment type, and COVID-19 (except for the maps on COVID-19). In general, there was a decrease in depression (−4.04%), anxiety (−2.68%), stress (−1.83%), COVID-19-related distress (−3.33%), and, to a lesser extent, loneliness (−0.28%) from baseline to 8-month survey. For most of the mental health variables, the baseline levels were higher in the northern part than the southern part of Sweden. In the eight monthly follow-up survey, there was a general decrease in all mental health measures and across the country. The northern-southern differences diminished but did not disappear. Regardless, the island of Gotland had the highest percentages in all mental health measures both at baseline and at 8-month follow-up.

### Evolution of mental health and sleep from baseline to eight months and from start to end of the study period

Figures S1 and S2 depict the evolution of the mental health indicators and sleep measures from baseline to 8-month follow-up survey. The level of stress was constant over time and above 40% of the total score. Loneliness was approximately 37% to 40%, increasing to its maximum at the 4-month follow-up and decreasing thereafter (Figure S1). The other mental health measures had values at the lower end, between 10% and 20%, with the total scores of depression, anxiety, and COVID-19-related distress increasing from baseline until the 3-month follow-up and decreasing thereafter. Finally, the sleep quantity index was around 10% and followed a pattern similar to COVID-19-related distress, depression, and anxiety. Figure S2 shows that the sleep quantity index increased from baseline until the 3-month follow-up indicating a deviation from normal sleep while sleep quality showed an opposite pattern with a lowest value (meaning poor sleep quality) at 3-month follow-up and a highest value at 6-month follow-up and on. In contrast, the mental health measures and sleep quantity decreased from 3-month follow-up until 8-month follow-up.

Figure 2 shows how levels of mental health symptomology and sleep measures varied throughout the study period. The levels of sleep quality were high in July 2020 and most of the participants had normal sleep quantity. A slight decreasing trend was seen until September 2020 in the scores of depression, anxiety, and COVID-19-related distress, whereas sleep quality began to drop sharply, and sleep quantity began to deviate from normal. From September to November 2020 there was an increase in levels of depression, anxiety, and COVID-19-related distress, with a peak noted in November-December 2020. Sleep quality was lowest in January 2021 and then increased steadily until August 2021. This correlated negatively with mental health measures and sleep quantity index which were lowest in July 2021. The temporal trend was less clear for stress, although a sharp decrease was observed from March to July 2021.

### Structural relationship between mental health and sleep at baseline, 4-months, and 8-months

Figure 3 represents the relationship between the mental health measures at three different time points (baseline, 4-month follow-up, and 8-month follow-up) and their correlations with COVID-19. There was a directional relationship between sleep quality and sleep quantity, namely that while the quality of sleep was highly influenced by sleep quantity, the other direction was weak. Sleep quality, on the other hand, was related to depression in such a way that depression affected sleep quality more than vice versa. Depression and anxiety were equally interrelated. Anxiety seemed to affect COVID-19-related distress more than how COVID-19-related distress affected anxiety. The relationship between anxiety and stress was similar. There was a weak relationship between stress and COVID-19-related distress. Loneliness was affected by depression, anxiety, and COVID-19-related distress. The correlations between mental health measures seemed to remain stable over time, however, the correlations between mental health measures and COVID-19 varied over time. At baseline the link to COVID-19 was stronger for depression, whereas at 8-month follow-up the link was stronger for COVID-19-related distress. Figure S3 illustrates these results after additional adjustment for sociodemographic factors, showing that sex, previous psychiatric disorder, and relationship status were also associated with mental health measures. In this figure, COVID-19 was connected to depression and age at baseline whereas only with age later.

### Relative risk of mental health burden over time based on sleep at baseline

Table 2 displays the risk of increasing mental health burden over time/at follow-up using as predictor sleep quality or quantity at baseline. A decreasing risk of depression, anxiety, COVID-19-related distress, and to a lesser extent also stress, was observed as sleep quality increased, regardless of multivariable adjustment. This risk decrease in mental health variables follows an approximately linear downward trend for all mental health outcomes. For example, the relative risk of depression is only 39.7 %, 31.9%, 20% and 11.5% for individuals with poor, medium, good, and very good sleep quality, respectively, compared to those with very poor sleep quality (models adjusted for COVID-19, sex, age, time, and recruitment type). Note that the relative risk of depression is significantly different for all five sleep categories. This holds true for anxiety and COVID-19 related distress as well.

**Table 2.** Relative risk (RR) of longitudinal depression, anxiety, Covid-19-related distress, and stress using as predictors sleep quality and sleep quantity index (SQI) at baseline after two different adjustments.

| Sleep variables at baseline | Depression |  | Anxiety |  | Covid-19 related distress |  | Stress |  |
| --- | --- | --- | --- | --- | --- | --- | --- | --- |
|  | *RR (95% CI)* | *p-value* | *RR (95% CI)* | *p-value* | *RR (95% CI)* | *p-value* | *RR (95% CI)* | *p-value* |
| **Model 1 Sleep Quality** |  |  |  |  |  |  |  |  |
| Sleep Quality (Very Poor) |  |  |  |  |  |  |  |  |
| Sleep Quality (Poor) | 0.66 (0.64-0.68) | 0.00e+00 | 0.66 (0.63-0.70) | 0.00e+00 | 0.77 (0.74-0.81) | 0.00e+00 | 0.96 (0.94-0.97) | 1.85e-07 |
| Sleep Quality (Medium) | 0.47 (0.45-0.49) | 0.00e+00 | 0.48 (0.45-0.51) | 0.00e+00 | 0.66 (0.63-0.69) | 0.00e+00 | 0.92 (0.90-0.93) | 0.00e+00 |
| Sleep Quality (Good) | 0.25 (0.24-0.26) | 0.00e+00 | 0.27 (0.26-0.29) | 0.00e+00 | 0.50 (0.48-0.53) | 0.00e+00 | 0.89 (0.88-0.91) | 0.00e+00 |
| Sleep Quality (Very Good) | 0.13 (0.12-0.14) | 0.00e+00 | 0.15 (0.14-0.16) | 0.00e+00 | 0.38 (0.35-0.40) | 0.00e+00 | 0.88 (0.86-0.90) | 0.00e+00 |
| **Model 2 Sleep Quality** |  |  |  |  |  |  |  |  |
| Sleep Quality (Very Poor) |  |  |  |  |  |  |  |  |
| Sleep Quality (Poor) | 0.70 (0.67-0.72) | 0.00e+00 | 0.70 (0.66-0.73) | 0.00e+00 | 0.80 (0.76-0.83) | 0.00e+00 | 0.96 (0.95-0.98) | 4.21e-06 |
| Sleep Quality (Medium) | 0.51 (0.49-0.53) | 0.00e+00 | 0.52 (0.49-0.55) | 0.00e+00 | 0.69 (0.66-0.72) | 0.00e+00 | 0.93 (0.91-0.94) | 0.00e+00 |
| Sleep Quality (Good) | 0.29 (0.28-0.30) | 0.00e+00 | 0.31 (0.29-0.33) | 0.00e+00 | 0.54 (0.51-0.56) | 0.00e+00 | 0.91 (0.89-0.92) | 0.00e+00 |
| Sleep Quality (Very Good) | 0.16 (0.15-0.17) | 0.00e+00 | 0.17 (0.16-0.19) | 0.00e+00 | 0.41 (0.38-0.43) | 0.00e+00 | 0.89 (0.88-0.91) | 0.00e+00 |
| **Model 1 Sleep Quantity Index** |  |  |  |  |  |  |  |  |
| SQI (7-9 hours) |  |  |  |  |  |  |  |  |
| SQI (6 or 10 hours) | 1.62 (1.58-1.66) | 0.0e+00 | 1.58 (1.53-1.63) | 0.00e+00 | 1.24 (1.21-1.27) | 0.00e+00 | 1.03 (1.02-1.04) | 1.01e-14 |
| SQI (5 or 11 hours) | 2.35 (2.26-2.44) | 0.0e+00 | 2.24 (2.14-2.34) | 0.00e+00 | 1.5 (1.44-1.55) | 0.00e+00 | 1.06 (1.05-1.07) | 0.00e+00 |
| SQI (5< or >11 hours) | 3.23 (3.04-3.43) | 0.0e+00 | 3.01 (2.78-3.25) | 0.00e+00 | 1.81 (1.70-1.93) | 0.00e+00 | 1.09 (1.06-1.12) | 9.12e-12 |
| **Model 2 Sleep Quantity Index** |  |  |  |  |  |  |  |  |
| SQI (7-9 hours) |  |  |  |  |  |  |  |  |
| SQI (6 or 10 hours) | 1.54 (1.51-1.58) | 0.00e+00 | 1.52 (1.47-1.57) | 0.00e+00 | 1.21 (1.18-1.24) | 0.00e+00 | 1.03 (1.02-1.03) | 2.71e-12 |
| SQI (5 or 11 hours) | 2.13 (2.05-2.21) | 0.00e+00 | 2.06 (1.97-2.15) | 0.00e+00 | 1.43 (1.38-1.49) | 0.00e+00 | 1.06 (1.04-1.07) | 0.00e+00 |
| SQI (5< or >11 hours) | 2.65 (2.50-2.82) | 0.00e+00 | 2.55 (2.36-2.75) | 0.00e+00 | 1.65 (1.55-1.76) | 0.00e+00 | 1.07 (1.05-1.10) | 2.90e-08 |

**Model 1:** Adjusted for COVID-19, sex, age, time & recruitment type
**Model 2:** Adjusted for COVID-19, sex, age, time, recruitment type, body mass index, habitual drinking, comorbidities, previous psychiatric disorder, relationship & current smoking

Conversely, an increasing risk of depression, anxiety, COVID-19-related distress, and to a lesser extent also stress, was noted as the sleep quantity index increased, regardless of the adjustments in the models. For example, the relative risk for depression increased linearly for each additional step diverging from normal sleep quantity (RR is 63.3%, 70.1% and 76.3%, for 6 or 10 hours, 5 or 11 hours and less than 5 or more than 11 hours, respectively (compared to 7-9 hours of sleep after adjustment for COVID-19, age, sex, time, and recruitment type). This pattern, with different risks ratios, was repeatedly seen in all the mental health variables in Table 2.

### Relative risk of worse sleep quality and quantity over time based on mental health indicators at baseline

Table 3 presents the relative risk of experiencing a certain level of sleep quality and the relative risk of deviation from a normal sleep quantity associated with the binary predictors of depression, anxiety, and COVID-19-related distress. High levels of mental health symptoms were associated with progressively declining risk of good sleep quality, meaning that high levels of mental health symptoms were strongly predictive of lower levels of sleep quality. This pattern occurs, with slightly different relative risks in the case high levels of anxiety and COVID-19 related distress, regardless of the adjustment and is very similar to the pattern seen in Table 2.

**Table 3.**
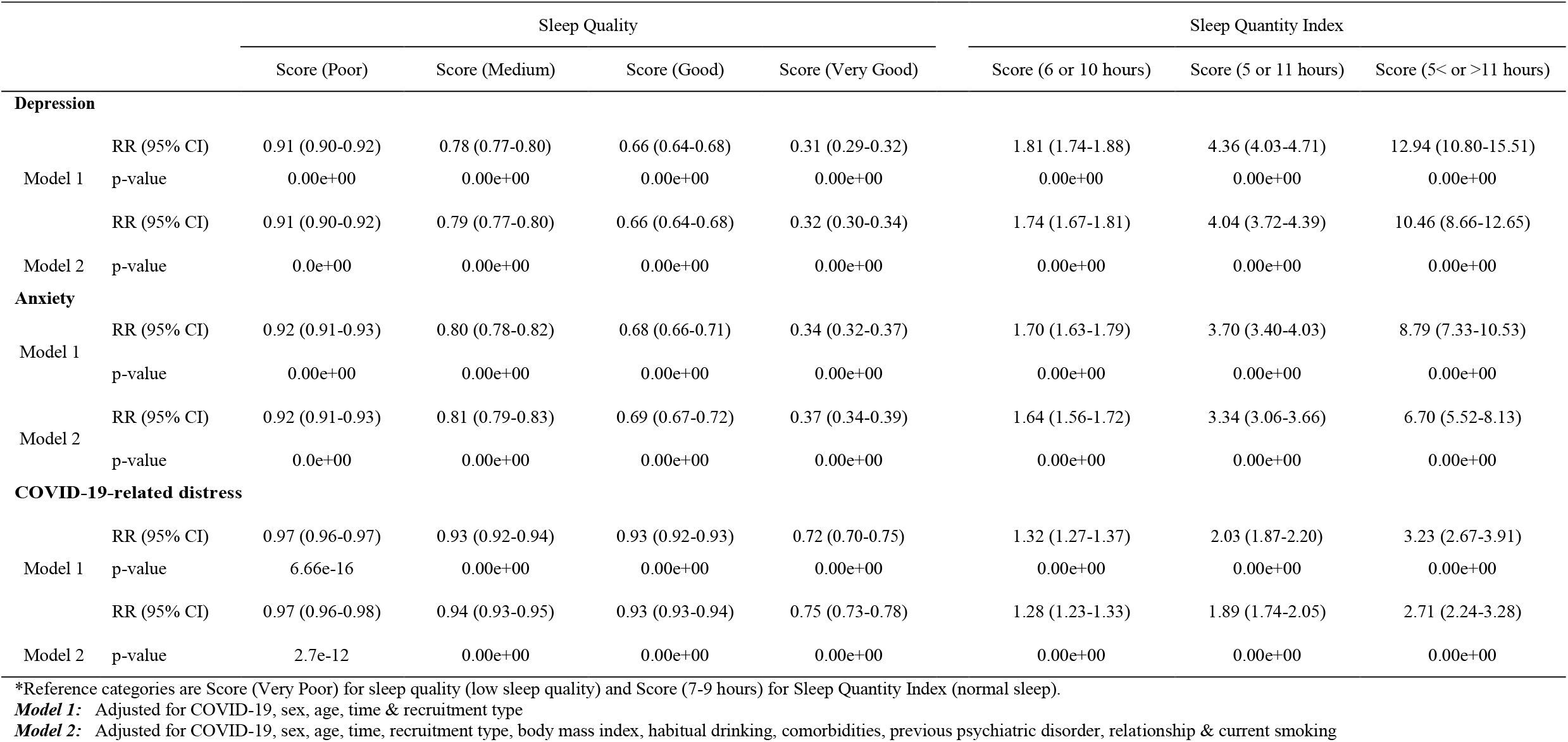
Relative Risk (RR) of longitudinal sleep quality and very poor sleep quantity index using as predictors depression, anxiety, and COVID-19-related distress at baseline after two different adjustments.

|  |  | Sleep Quality |  |  |  | Sleep Quantity Index |  |  |
| --- | --- | --- | --- | --- | --- | --- | --- | --- |
|  |  | Score (Poor) | Score (Medium) | Score (Good) | Score (Very Good) | Score (6 or 10 hours) | Score (5 or 11 hours) | Score (5< or >11 hours) |
| **Depression** |  |  |  |  |  |  |  |  |
| Model 1 | RR (95% CI) | 0.91 (0.90-0.92) | 0.78 (0.77-0.80) | 0.66 (0.64-0.68) | 0.31 (0.29-0.32) | 1.81 (1.74-1.88) | 4.36 (4.03-4.71) | 12.94 (10.80-15.51) |
|  | p-value | 0.00e+00 | 0.00e+00 | 0.00e+00 | 0.00e+00 | 0.00e+00 | 0.00e+00 | 0.00e+00 |
| Model 2 | RR (95% CI) | 0.91 (0.90-0.92) | 0.79 (0.77-0.80) | 0.66 (0.64-0.68) | 0.32 (0.30-0.34) | 1.74 (1.67-1.81) | 4.04 (3.72-4.39) | 10.46 (8.66-12.65) |
|  | p-value | 0.0e+00 | 0.00e+00 | 0.00e+00 | 0.00e+00 | 0.00e+00 | 0.00e+00 | 0.00e+00 |
| **Anxiety** |  |  |  |  |  |  |  |  |
| Model 1 | RR (95% CI) | 0.92 (0.91-0.93) | 0.80 (0.78-0.82) | 0.68 (0.66-0.71) | 0.34 (0.32-0.37) | 1.70 (1.63-1.79) | 3.70 (3.40-4.03) | 8.79 (7.33-10.53) |
|  | p-value | 0.00e+00 | 0.00e+00 | 0.00e+00 | 0.00e+00 | 0.00e+00 | 0.00e+00 | 0.00e+00 |
| Model 2 | RR (95% CI) | 0.92 (0.91-0.93) | 0.81 (0.79-0.83) | 0.69 (0.67-0.72) | 0.37 (0.34-0.39) | 1.64 (1.56-1.72) | 3.34 (3.06-3.66) | 6.70 (5.52-8.13) |
|  | p-value | 0.0e+00 | 0.00e+00 | 0.00e+00 | 0.00e+00 | 0.00e+00 | 0.00e+00 | 0.00e+00 |
| **COVID-19-related distress** |  |  |  |  |  |  |  |  |
| Model 1 | RR (95% CI) | 0.97 (0.96-0.97) | 0.93 (0.92-0.94) | 0.93 (0.92-0.93) | 0.72 (0.70-0.75) | 1.32 (1.27-1.37) | 2.03 (1.87-2.20) | 3.23 (2.67-3.91) |
|  | p-value | 6.66e-16 | 0.00e+00 | 0.00e+00 | 0.00e+00 | 0.00e+00 | 0.00e+00 | 0.00e+00 |
| Model 2 | RR (95% CI) | 0.97 (0.96-0.98) | 0.94 (0.93-0.95) | 0.93 (0.93-0.94) | 0.75 (0.73-0.78) | 1.28 (1.23-1.33) | 1.89 (1.74-2.05) | 2.71 (2.24-3.28) |
|  | p-value | 2.7e-12 | 0.00e+00 | 0.00e+00 | 0.00e+00 | 0.00e+00 | 0.00e+00 | 0.00e+00 |

\*Reference categories are Score (Very Poor) for sleep quality (low sleep quality) and Score (7-9 hours) for Sleep Quantity Index (normal sleep).
**Model 1:** Adjusted for COVID-19, sex, age, time & recruitment type
**Model 2:** Adjusted for COVID-19, sex, age, time, recruitment type, body mass index, habitual drinking, comorbidities, previous psychiatric disorder, relationship & current smoking

On the other hand, the relationship between mental health and the divergence from normal sleep quantity is quadratic in the case of depression and anxiety. For example, the relative risk to diverge from normal sleep hours when experiencing high depressive symptoms is 1.81, 4.36 and 12.94 (6 or 10 hours, 5 or 11 hours and less than 5 or more than 11 hours, respectively (after adjustment for COVID-19, age, sex, time, and recruitment type)). In the case of experiencing COVID-19-related distress this relationship is more-or-less linear, for instance the relative risk to diverge from normal sleep hours after adjustment for COVID-19, age, sex, and recruitment type is 1.32, 2.03 and 3.23 (6 or 10 hours, 5 or 11 hours and less than 5 or more than 11 hours, respectively).

## Discussion

This study aimed to investigate the longitudinal link between sleep quality and quantity to several mental health measures during the current COVID-19 pandemic. Our results show a strong bidirectional relationship between sleep disturbance and depression, anxiety, and COVID-19-related distress. Such relationship was stable over time and followed to some extent the COVID-19 incidence, consistent with previous studies (Halsøy, Johnson, Hoffart, & Ebrahimi, 2021). This study began in July 2020. Although we did not study the first wave of pandemic in Sweden, we collected monthly data during the second and third waves, when mitigating measures were stricter in the country.

### Bidirectional relationship between sleep and mental health

The bidirectionality of the relationship between anxiety, depression, and sleep disturbance found in our study is consistent with the previous literature (Alvaro, Roberts, & Harris, 2013). Kalmbach et al. (2018) suggesting that similar bidirectional mechanisms are at play during pandemic times. Highly stressful societal events, like the COVID-19 pandemic, could indeed be a trigger for sleep problems which could subsequently give rise to symptoms of depression and anxiety. Mental health burden and sleep problems can continue even when the triggering event has passed (Morin et al., 2009). Several studies have shown that the adoption of measures to hamper the spread of COVID-19 during 2020 (Ahmed et al., 2020) and the fear about the virus are important triggers of stress, leading to subsequent poor sleep, depression, and anxiety (Siddique, Ahmed, & Hossain, 2021).

Even though there never was a strict lockdown in Sweden, other measures were adopted such as working from home whenever possible, participant limits to private and public events, and event cancellations. There was an increased incidence of cases during September 2020 leading to the adoption of more restrictive measures from end of October of 2020 until April 2021, when the most vulnerable individuals of the population were vaccinated. Thereafter, the restrictions were gradually lifted and eliminated in July 2021. Our results follow this pattern to some extent. Despite the high prevalence of seasonal affective disorder during the winter in Sweden (Rastad, Sjöden, & Ulfberg, 2005), the effect of the pandemic is also significant, explaining partly the geographic variation of the mental health measures noted in the study. Nevertheless, compared with the pre-pandemic levels, there was an increase in depression, anxiety, and COVID-19-related distress already at the baseline of the Omtanke2020 study (Lovik et al., 2021).

### Mental health, sleep, and COVID-19

At baseline of the present study, only 5.57% of participants had had COVID-19. The level of COVID-19-related distress was however much higher at baseline, compared with the 8-month follow-up when 42.42% of the participants had contracted the disease. There was also a negative correlation, although weak, between COVID-19-related distress and COVID-19. On the other hand, there was no correlation between COVID-19 and sleep quality or quantity. We hypothesize that people with the greatest fear for the virus and those not infected were likely individuals with higher levels of mental health burden and poor sleep. The COVID-19-related distress questionnaire is a modification of the PC-PTSD-5 and may reflect the fear of contagion and not only the traumatic event of having had the virus. That could be the reason why people who had a mild illness showed a lower level of COVID-19-related distress than those who had not been infected (Magnúsdóttir et al., 2022).

### Strengths and weaknesses

The large sample size is a strength of this study. Among the 9,035 individuals included in the study, 3,883 had contracted COVID-19. The second strength is the temporal resolution, as we had both baseline survey and eight monthly follow-up surveys covering the second and third epidemic waves as well as the start of vaccination in Sweden.

Our study also has limitations. On the one hand, as participation was open to all, our sample is not representative of the general Swedish population. For example, 81% of the participants were women and there was a relatively low proportion of younger people. Selection bias due to the method of recruitment could therefore not be excluded. For instance, it has been shown that people recruited through social media campaigns had higher levels of anxiety and depression, compared with people recruited by invitation, in the Omtanke2020 study (Lovik et al., 2021). To consider these limitations, we adjusted in all analyses for sex, age, and recruitment type. The modification of the PC-PTSD-5 scale to make it specific for the pandemic is another concern as it included three items on thinking and dreaming about COVID-19. As a result, this scale might have evaluated the post-traumatic stress of being in a pandemic rather than contracting the disease. We therefore referred to this outcome as COVID-19-related distress throughout this paper. Finally, the low correlation noted between stress and other mental health measures may be partly due to the use of the abbreviated form of the PSS test, with only 4 items instead of 10, which has not been validated in Sweden and shows relatively poor psychometric properties in our sample.

## Conclusion

In summary, in this study, we found that there was a strong bi-directional link between mental health and sleep during the current COVID-19 pandemic in Sweden, and that the temporal pattern of these measures was also correlated to the burden of the pandemic.

## Supporting information

All supplementary figures and tables

## Data Availability

Data produced in the present study are available upon reasonable request to the authors

## Acknowledgement

This study was supported by NordForsk (project No. 105668) and Horizon2020 (CoMorMent, 847776). The authors also thank the participants of the Omtanke2020 study.

## Conflict of Interest

The authors declare no conflict of interest.

## Ethical approval

The study was approved (DNR 2020-01785) by the Swedish Ethical Review Authority on 3 June 2020.

## Notes

### Competing Interest Statement

The authors have declared no competing interest.

### Author Declarations

The Omtanke2020 study has been registered and approved (DNR 2020-01785) by Etikprovningsmyndigheten (Swedish National Ethics Board) on 3 June 2020.

