## Supplementary material for "Unravelling the link between sleep and mental health during the COVID-19 pandemic": All supplementary figures and tables

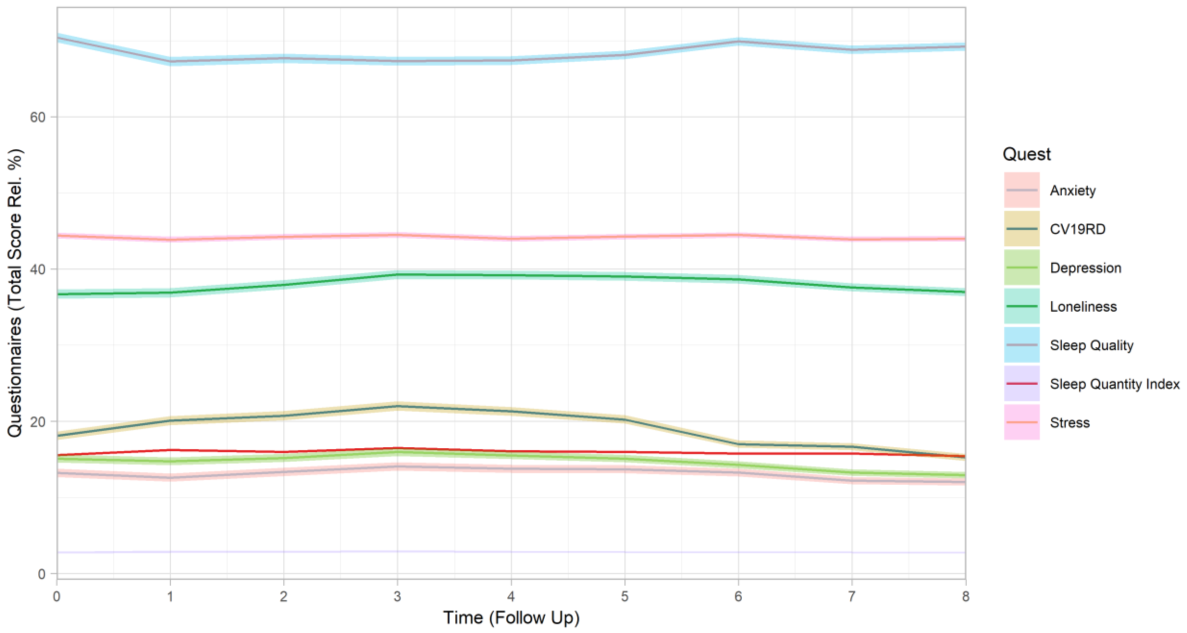


*Figure S 1 Temporal pattern (relative percentage of total score) of depression, anxiety, COVID-19-related distress (CV19RD), stress, loneliness, sleep quality, and sleep quantity index, adjusted for age, sex, recruitment type and COVID-19.*

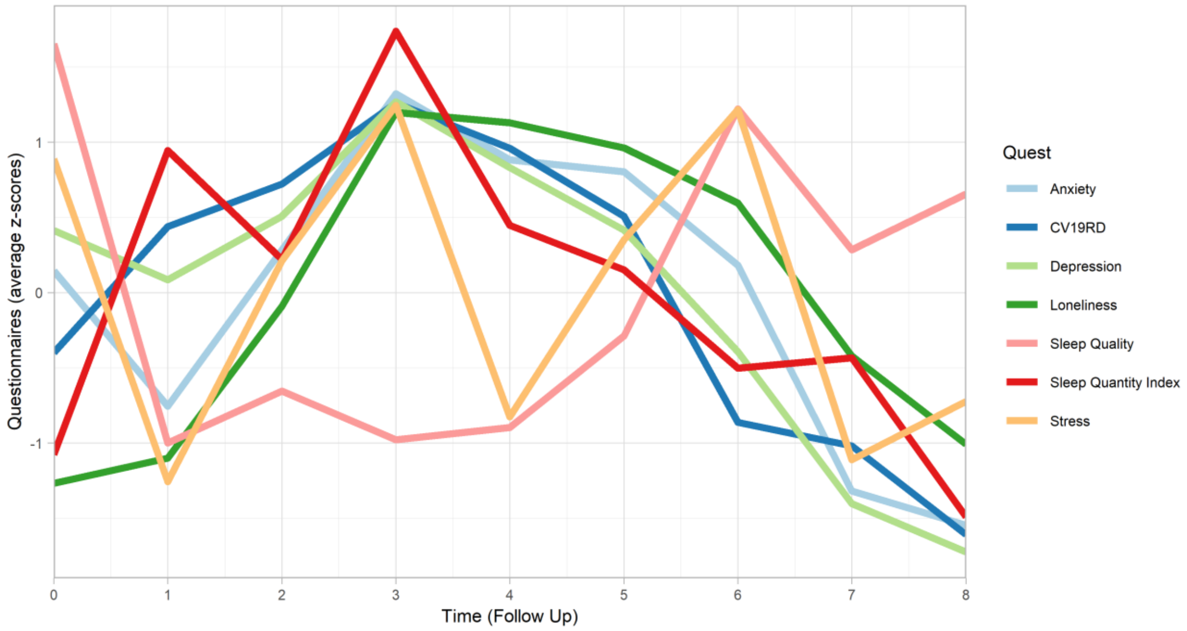


*Figure S 2.- Follow Up variation (average Z-score) of Anxiety, COVID-19-related Distress (CV19RD), Depression, Loneliness, Sleep Hour Index, Sleep Quality and Stress. Adjusted for Age, Sex, Recruitment type and COVID-19 status.*

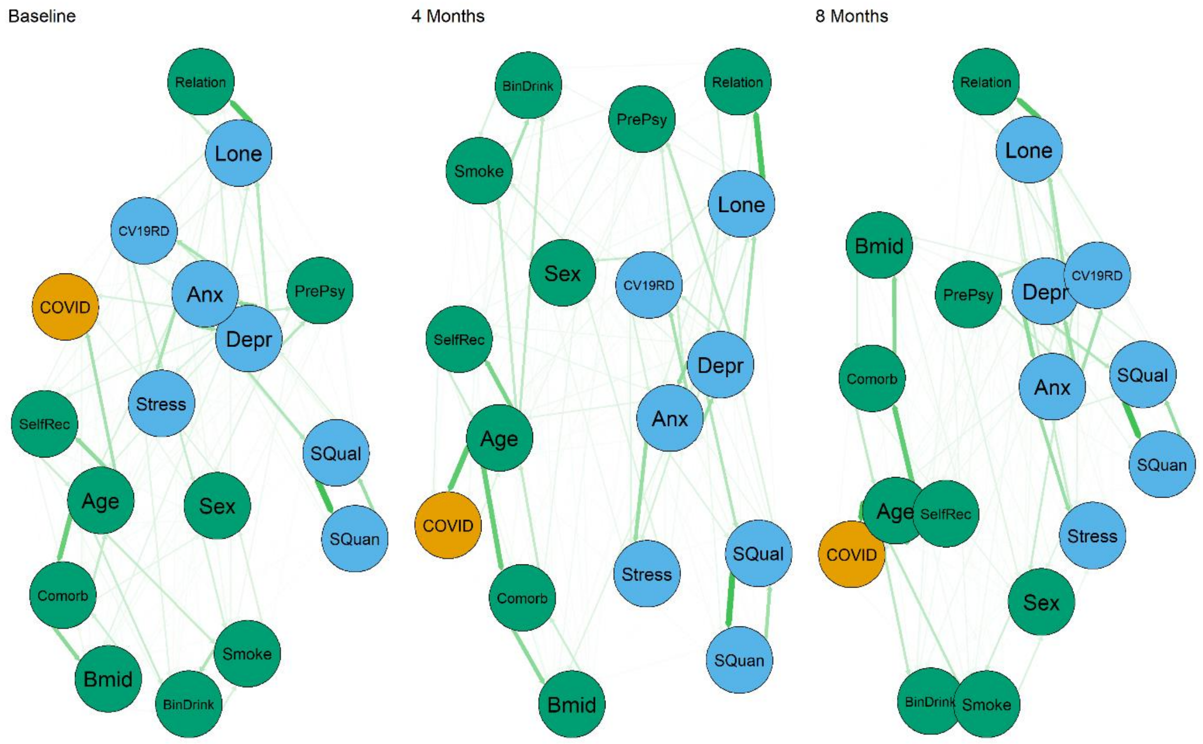


*Figure S 3. Relationships between mental health, in green (Depression (Depr), Anxiety (Anx), COVID-19-related Distress (CV19RD), stress, loneliness (Lone), sleep quality (SQual) and sleep quantity (SQuan), sociodemographic, in blue (sex, age, recruitment type (SelfRec), relationship (Relation), previous psychiatric diagnosis (PrePsy), body mass index (BMID), habitual drinking (BinDrink), smoking and comorbidities (Comorb)) and COVID-19 status (COVID) variables.*

| | Depression | | Anxiety | | CV19RD | | Stress | |
|---|---|---|---|---|---|---|---|---|
| | *RR (95% CI)* | *p-value* | *RR (95% CI)* | *p-value* | *RR (95% CI)* | *p-value* | *RR (95% CI)* | *p-value* |
| **Model 1 Sleep Quality** | | | | | | | | |
| (Intercept) | 19.78 (18.52-21.12) | 0.00e+00 | 17.92 (16.54-19.43) | 0.00e+00 | 6.83 (6.36-7.34) | 0.00e+00 | 8.75 (8.55-8.95) | 0.00e+00 |
| Sleep Quality (Poor) | 0.66 (0.64-0.68) | 0.00e+00 | 0.66 (0.63-0.7) | 0.00e+00 | 0.77 (0.74-0.81) | 0.00e+00 | 0.96 (0.94-0.97) | 1.85e-07 |
| Sleep Quality (Medium) | 0.47 (0.45-0.49) | 0.00e+00 | 0.48 (0.45-0.51) | 0.00e+00 | 0.66 (0.63-0.69) | 0.00e+00 | 0.92 (0.9-0.93) | 0.00e+00 |
| Sleep Quality (Good) | 0.25 (0.24-0.26) | 0.00e+00 | 0.27 (0.26-0.29) | 0.00e+00 | 0.5 (0.48-0.53) | 0.00e+00 | 0.89 (0.88-0.91) | 0.00e+00 |
| Sleep Quality (Very Good) | 0.13 (0.12-0.14) | 0.00e+00 | 0.15 (0.14-0.16) | 0.00e+00 | 0.38 (0.35-0.4) | 0.00e+00 | 0.88 (0.86-0.9) | 0.00e+00 |
| COVID-19 | 0.99 (0.96-1.02) | 3.81e-01 | 0.95 (0.92-0.99) | 1.06e-02 | 0.92 (0.89-0.94) | 3.11e-08 | 0.98 (0.97-0.99) | 6.52e-05 |
| Time | 0.98 (0.98-0.99) | 0.00e+00 | 0.99 (0.99-1) | 1.11e-04 | 0.97 (0.97-0.98) | 0.00e+00 | 1 (1-1) | 4.60e-01 |
| Sex (Male) | 0.79 (0.76-0.83) | 0.00e+00 | 0.73 (0.69-0.77) | 0.00e+00 | 0.71 (0.68-0.75) | 0.00e+00 | 0.92 (0.91-0.93) | 0.00e+00 |
| Age | 0.99 (0.99-0.99) | 0.00e+00 | 0.98 (0.98-0.99) | 0.00e+00 | 1 (1-1) | 9.14e-10 | 1 (1-1) | 0.00e+00 |
| Recruitment Type | 1.28 (1.24-1.32) | 0.00e+00 | 1.29 (1.24-1.34) | 0.00e+00 | 1.21 (1.18-1.25) | 0.00e+00 | 1.03 (1.02-1.04) | 3.81e-12 |
| **Model 2 Sleep Quality** | | | | | | | | |
| (Intercept) | 17.27 (16.12-18.5) | 0.00e+00 | 13.41 (12.27-14.66) | 0.00e+00 | 6.3 (5.82-6.82) | 0.00e+00 | 8.48 (8.27-8.68) | 0.00e+00 |
| Sleep Quality (Poor) | 0.7 (0.67-0.72) | 0.00e+00 | 0.7 (0.66-0.73) | 0.00e+00 | 0.8 (0.76-0.83) | 0.00e+00 | 0.96 (0.95-0.98) | 4.21e-06 |
| Sleep Quality (Medium) | 0.51 (0.49-0.53) | 0.00e+00 | 0.52 (0.49-0.55) | 0.00e+00 | 0.69 (0.66-0.72) | 0.00e+00 | 0.93 (0.91-0.94) | 0.00e+00 |
| Sleep Quality (Good) | 0.29 (0.28-0.3) | 0.00e+00 | 0.31 (0.29-0.33) | 0.00e+00 | 0.54 (0.51-0.56) | 0.00e+00 | 0.91 (0.89-0.92) | 0.00e+00 |
| Sleep Quality (Very Good) | 0.16 (0.15-0.17) | 0.00e+00 | 0.17 (0.16-0.19) | 0.00e+00 | 0.41 (0.38-0.43) | 0.00e+00 | 0.89 (0.88-0.91) | 0.00e+00 |
| COVID-19 | 0.99 (0.96-1.02) | 3.86e-01 | 0.95 (0.92-0.99) | 9.72e-03 | 0.92 (0.89-0.95) | 4.44e-08 | 0.98 (0.97-0.99) | 7.77e-05 |
| Time | 0.98 (0.98-0.99) | 0.00e+00 | 0.99 (0.99-1) | 1.01e-04 | 0.97 (0.97-0.98) | 0.00e+00 | 1 (1-1) | 4.35e-01 |
| Sex (Male) | 0.83 (0.79-0.86) | 0.00e+00 | 0.77 (0.72-0.81) | 0.00e+00 | 0.72 (0.69-0.75) | 0.00e+00 | 0.93 (0.91-0.94) | 0.00e+00 |
| Age | 0.99 (0.99-0.99) | 0.00e+00 | 0.98 (0.98-0.99) | 0.00e+00 | 1 (1-1) | 9.91e-06 | 1 (1-1) | 0.00e+00 |
| Recruitment Type | 1.17 (1.14-1.21) | 0.00e+00 | 1.2 (1.16-1.25) | 0.00e+00 | 1.17 (1.13-1.2) | 0.00e+00 | 1.02 (1.01-1.03) | 1.75e-07 |
| BMID (obese) | 1.13 (1.08-1.17) | 2.78e-09 | 0.96 (0.91-1.01) | 9.98e-02 | 1 (0.96-1.05) | 8.77e-01 | 0.98 (0.97-1) | 2.27e-02 |
| BMID (overweight) | 1.02 (0.99-1.06) | 1.36e-01 | 0.96 (0.92-1) | 3.26e-02 | 1 (0.97-1.03) | 9.58e-01 | 0.99 (0.98-1) | 2.31e-02 |
| Habitual Drinking | 0.97 (0.94-1.01) | 1.04e-01 | 0.98 (0.94-1.02) | 3.64e-01 | 1.04 (1.01-1.07) | 2.12e-02 | 0.98 (0.97-0.99) | 5.89e-05 |
| Comorbidity (1) | 1.11 (1.07-1.14) | 1.05e-09 | 1.11 (1.07-1.16) | 1.16e-06 | 1.11 (1.08-1.15) | 3.44e-10 | 1 (0.99-1.01) | 5.63e-01 |
| Comorbidity (2) | 1.15 (1.09-1.21) | 1.01e-07 | 1.19 (1.11-1.28) | 2.43e-06 | 1.24 (1.17-1.3) | 2.33e-15 | 1.02 (1-1.04) | 3.36e-02 |
| Comorbidity (3+) | 1.3 (1.19-1.42) | 3.30e-09 | 1.27 (1.12-1.43) | 2.36e-04 | 1.13 (1.04-1.23) | 4.34e-03 | 1.02 (0.99-1.04) | 2.38e-01 |
| Psychiatric Diag | 1.45 (1.41-1.49) | 0.00e+00 | 1.56 (1.5-1.62) | 0.00e+00 | 1.19 (1.16-1.23) | 0.00e+00 | 1.07 (1.06-1.08) | 0.00e+00 |
| Relationship | 0.81 (0.79-0.83) | 0.00e+00 | 0.99 (0.95-1.03) | 5.21e-01 | 0.95 (0.92-0.98) | 3.61e-04 | 1 (0.99-1.01) | 8.17e-01 |
| Smoking | 1.07 (1.03-1.11) | 6.25e-04 | 1.05 (1-1.11) | 3.24e-02 | 1.06 (1.02-1.1) | 3.84e-03 | 1 (0.98-1.01) | 4.85e-01 |
| **Model 1 Sleep Quant Index** | | | | | | | | |
| (Intercept) | 6.53 (6.1-6.99) | 0.0e+00 | 6.2 (5.73-6.71) | 0.00e+00 | 3.79 (3.56-4.04) | 0.00e+00 | 7.93 (7.8-8.07) | 0.00e+00 |
| SQI (6 or 10 hours) | 1.62 (1.58-1.66) | 0.0e+00 | 1.58 (1.53-1.63) | 0.00e+00 | 1.24 (1.21-1.27) | 0.00e+00 | 1.03 (1.02-1.04) | 1.01e-14 |
| SQI (5 or 11 hours) | 2.35 (2.26-2.44) | 0.0e+00 | 2.24 (2.14-2.34) | 0.00e+00 | 1.5 (1.44-1.55) | 0.00e+00 | 1.06 (1.05-1.07) | 0.00e+00 |
| SQI (5< or >11 hours) | 3.23 (3.04-3.43) | 0.0e+00 | 3.01 (2.78-3.25) | 0.00e+00 | 1.81 (1.7-1.93) | 0.00e+00 | 1.09 (1.06-1.12) | 9.12e-12 |
| COVID-19 | 1 (0.97-1.04) | 8.6e-01 | 0.97 (0.93-1.01) | 1.02e-01 | 0.92 (0.89-0.95) | 1.20e-06 | 0.98 (0.97-0.99) | 2.03e-04 |
| Time | 0.98 (0.98-0.99) | 0.0e+00 | 0.99 (0.99-1) | 2.97e-06 | 0.97 (0.97-0.98) | 0.00e+00 | 1 (1-1) | 3.35e-01 |
| Sex (Male) | 0.75 (0.71-0.79) | 0.0e+00 | 0.69 (0.65-0.74) | 0.00e+00 | 0.69 (0.66-0.72) | 0.00e+00 | 0.91 (0.9-0.93) | 0.00e+00 |
| Age | 0.99 (0.99-0.99) | 0.0e+00 | 0.98 (0.98-0.98) | 0.00e+00 | 1 (1-1) | 4.13e-05 | 1 (1-1) | 0.00e+00 |
| Recruitment Type | 1.34 (1.29-1.38) | 0.0e+00 | 1.35 (1.29-1.4) | 0.00e+00 | 1.24 (1.2-1.28) | 0.00e+00 | 1.03 (1.02-1.04) | 3.36e-14 |
| **Model 2 Sleep Quant Index** | | | | | | | | |
| (Intercept) | 6.13 (5.71-6.59) | 0.00e+00 | 4.94 (4.54-5.39) | 0.00e+00 | 3.64 (3.39-3.91) | 0.00e+00 | 7.76 (7.61-7.9) | 0.00e+00 |
| SQI (6 or 10 hours) | 1.54 (1.51-1.58) | 0.00e+00 | 1.52 (1.47-1.57) | 0.00e+00 | 1.21 (1.18-1.24) | 0.00e+00 | 1.03 (1.02-1.03) | 2.71e-12 |
| SQI (5 or 11 hours) | 2.13 (2.05-2.21) | 0.00e+00 | 2.06 (1.97-2.15) | 0.00e+00 | 1.43 (1.38-1.49) | 0.00e+00 | 1.06 (1.04-1.07) | 0.00e+00 |
| SQI (5< or >11 hours) | 2.65 (2.5-2.82) | 0.00e+00 | 2.55 (2.36-2.75) | 0.00e+00 | 1.65 (1.55-1.76) | 0.00e+00 | 1.07 (1.05-1.1) | 2.90e-08 |
| COVID-19 | 1 (0.97-1.04) | 7.83e-01 | 0.97 (0.93-1.01) | 1.08e-01 | 0.92 (0.9-0.95) | 1.33e-06 | 0.98 (0.97-0.99) | 1.94e-04 |
| Time | 0.98 (0.98-0.99) | 0.00e+00 | 0.99 (0.99-1) | 2.04e-06 | 0.97 (0.97-0.98) | 0.00e+00 | 1 (1-1) | 3.35e-01 |
| Sex (Male) | 0.79 (0.76-0.83) | 0.00e+00 | 0.74 (0.69-0.78) | 0.00e+00 | 0.7 (0.67-0.74) | 0.00e+00 | 0.92 (0.91-0.93) | 0.00e+00 |
| Age | 0.99 (0.99-0.99) | 0.00e+00 | 0.98 (0.98-0.98) | 0.00e+00 | 1 (1-1) | 6.48e-03 | 1 (1-1) | 0.00e+00 |
| Recruitment Type | 1.2 (1.16-1.24) | 0.00e+00 | 1.23 (1.18-1.28) | 0.00e+00 | 1.18 (1.14-1.21) | 0.00e+00 | 1.02 (1.02-1.03) | 2.80e-08 |
| BMID (obese) | 1.12 (1.08-1.18) | 3.20e-07 | 0.95 (0.9-1.01) | 1.15e-01 | 1.01 (0.96-1.05) | 8.16e-01 | 0.99 (0.97-1) | 2.89e-02 |
| BMID (overweight) | 1.03 (0.99-1.06) | 1.42e-01 | 0.96 (0.92-1) | 6.04e-02 | 1 (0.97-1.03) | 9.63e-01 | 0.99 (0.98-1) | 2.68e-02 |
| Habitual Drinking | 0.97 (0.93-1) | 7.25e-02 | 0.97 (0.93-1.02) | 2.47e-01 | 1.04 (1-1.07) | 3.44e-02 | 0.98 (0.97-0.99) | 6.70e-05 |
| Comorbidity (1) | 1.14 (1.1-1.18) | 5.69e-12 | 1.14 (1.09-1.2) | 9.34e-09 | 1.13 (1.09-1.17) | 8.28e-12 | 1 (0.99-1.01) | 3.87e-01 |
| Comorbidity (2) | 1.21 (1.13-1.28) | 5.74e-09 | 1.25 (1.15-1.35) | 8.58e-08 | 1.27 (1.2-1.34) | 0.00e+00 | 1.02 (1-1.04) | 1.34e-02 |
| Comorbidity (3+) | 1.37 (1.25-1.5) | 3.62e-11 | 1.33 (1.17-1.51) | 1.13e-05 | 1.17 (1.07-1.27) | 5.28e-04 | 1.02 (0.99-1.05) | 1.52e-01 |
| Psychiatric Diag. | 1.6 (1.55-1.66) | 0.00e+00 | 1.71 (1.64-1.78) | 0.00e+00 | 1.25 (1.21-1.29) | 0.00e+00 | 1.07 (1.06-1.08) | 0.00e+00 |
| Relationship | 0.81 (0.79-0.84) | 0.00e+00 | 0.99 (0.95-1.03) | 5.77e-01 | 0.95 (0.92-0.98) | 6.67e-04 | 1 (0.99-1.01) | 7.86e-01 |
| Smoking | 1.08 (1.04-1.13) | 2.93e-04 | 1.07 (1.02-1.13) | 1.16e-02 | 1.07 (1.02-1.11) | 1.83e-03 | 1 (0.98-1.01) | 6.05e-01 |

***Model 1:*** Adjusted COVID-19, Sex, Time & Recruitment Type

***Model 2:*** Adjusted COVID-19, Sex, Time, Recruitment Type, BMID, Drinking, Comorbidities, Previous Psychiatric Diag., Relationship & Smoking

*Table S 1. Complete table of Relative Risk (RR) of depression, anxiety, Covid-19-related distress, and stress using as predictors sleep quality and sleep quantity index using two different adjustments.*

| | Sleep Quality | | | | | | | | Sleep Quantity Index | | | | | |
|---|---|---|---|---|---|---|---|---|---|---|---|---|---|---|
| | Score (2) | | Score (3) | | Score (4) | | Score (5) | | Score (1) | | Score (2) | | Score (3) | |
| | *RR (95% CI)* | *p-value* | *RR (95% CI)* | *p-value* | *RR (95% CI)* | *p-value* | *RR (95% CI)* | *p-value* | *RR (95% CI)* | *p-value* | *RR (95% CI)* | *p-value* | *RR (95% CI)* | *p-value* |
| **Depression** | | | | | | | | | | | | | | |
| (Intercept) | 2 (1.95-2.04) | 0.0e+00 | 3.02 (2.94-3.1) | 0.0e+00 | 4.04 (3.98-4.1) | 0.0e+00 | 4.76 (4.59-4.93) | 0.0e+00 | 0.21 (0.19-0.23) | 0.0e+00 | 0.08 (0.07-0.1) | 0.0e+00 | 0.01 (0.01-0.02) | 0.0e+00 |
| Depression | 0.91 (0.9-0.92) | 0.0e+00 | 0.78 (0.77-0.8) | 0.0e+00 | 0.66 (0.64-0.68) | 0.0e+00 | 0.31 (0.29-0.32) | 0.0e+00 | 1.81 (1.74-1.88) | 0.0e+00 | 4.36 (4.03-4.71) | 0.0e+00 | 12.94 (10.8-15.51) | 0.0e+00 |
| COVID-19 | 0.99 (0.98-1) | 1.4e-01 | 0.99 (0.97-1) | 9.5e-02 | 0.99 (0.98-1) | 5.1e-02 | 0.98 (0.96-1) | 2.4e-02 | 1.07 (1.02-1.12) | 6.5e-03 | 1.05 (0.96-1.16) | 2.9e-01 | 1.14 (0.94-1.39) | 1.7e-01 |
| Time | 1 (1-1) | 1.1e-03 | 1 (1-1) | 3.0e-02 | 1 (1-1) | 2.4e-04 | 1 (1-1) | 1.3e-03 | 1 (0.99-1) | 1.1e-01 | 1 (0.99-1.01) | 9.7e-01 | 1.03 (1.01-1.05) | 6.7e-03 |
| Sex (Male) | 1 (0.98-1.01) | 5.8e-01 | 1.01 (0.99-1.02) | 3.9e-01 | 1.01 (1-1.01) | 2.1e-01 | 1.03 (1.01-1.05) | 3.1e-03 | 0.96 (0.9-1.03) | 2.4e-01 | 0.98 (0.85-1.12) | 7.3e-01 | 1.07 (0.8-1.43) | 6.6e-01 |
| Age | 1 (1-1) | 1.2e-03 | 1 (1-1) | 1.4e-03 | 1 (1-1) | 1.7e-04 | 1 (1-1) | 5.5e-01 | 1 (1-1.01) | 1.6e-10 | 1.01 (1.01-1.01) | 4.8e-13 | 1.01 (1.01-1.02) | 3.2e-07 |
| Recruitment type* | 1 (0.99-1.01) | 4.2e-01 | 0.99 (0.97-1) | 6.3e-02 | 1 (0.99-1) | 2.9e-01 | 0.98 (0.97-1) | 1.0e-01 | 1 (0.96-1.05) | 8.9e-01 | 1.1 (1-1.21) | 5.1e-02 | 1.26 (1.03-1.56) | 2.7e-02 |
| **Anxiety** | | | | | | | | | | | | | | |
| (Intercept) | 1.96 (1.92-2) | 0.0e+00 | 2.91 (2.83-2.99) | 0.0e+00 | 3.99 (3.92-4.05) | 0.0e+00 | 4.47 (4.26-4.69) | 0.0e+00 | 0.22 (0.2-0.24) | 0.0e+00 | 0.1 (0.08-0.12) | 0.0e+00 | 0.02 (0.01-0.03) | 0.0e+00 |
| Anxiety | 0.92 (0.91-0.93) | 0.0e+00 | 0.8 (0.78-0.82) | 0.0e+00 | 0.68 (0.66-0.71) | 0.0e+00 | 0.34 (0.32-0.36) | 0.0e+00 | 1.7 (1.63-1.78) | 0.0e+00 | 3.7 (3.4-4.03) | 0.0e+00 | 8.79 (7.33-10.53) | 0.0e+00 |
| COVID-19 | 0.99 (0.98-1) | 1.1e-01 | 0.98 (0.97-1) | 3.1e-02 | 0.99 (0.98-1) | 9.3e-03 | 0.97 (0.94-0.99) | 7.5e-03 | 1.07 (1.02-1.13) | 3.4e-03 | 1.08 (0.98-1.19) | 1.2e-01 | 1.2 (0.98-1.46) | 7.6e-02 |
| Time | 1 (1-1) | 2.7e-02 | 1 (1-1) | 3.9e-01 | 1 (1-1) | 7.1e-02 | 1 (1-1) | 3.8e-01 | 0.99 (0.99-1) | 1.6e-02 | 0.99 (0.99-1) | 2.6e-01 | 1.02 (1-1.04) | 6.6e-02 |
| Sex (Male) | 1 (0.99-1.01) | 9.8e-01 | 1.01 (1-1.03) | 1.1e-01 | 1.01 (1-1.02) | 4.1e-02 | 1.05 (1.02-1.08) | 1.8e-04 | 0.95 (0.88-1.01) | 1.1e-01 | 0.93 (0.81-1.07) | 3.3e-01 | 0.98 (0.73-1.33) | 9.1e-01 |
| Age | 1 (1-1) | 3.6e-03 | 1 (1-1) | 4.8e-02 | 1 (1-1) | 9.6e-04 | 1 (1-1) | 6.9e-01 | 1 (1-1.01) | 4.3e-09 | 1.01 (1.01-1.01) | 1.7e-11 | 1.01 (1.01-1.02) | 2.3e-06 |
| Recruitment type* | 0.99 (0.98-1) | 8.5e-02 | 0.98 (0.96-0.99) | 1.7e-03 | 0.99 (0.98-1) | 5.4e-03 | 0.96 (0.93-0.98) | 3.5e-04 | 1.02 (0.98-1.08) | 3.2e-01 | 1.17 (1.06-1.29) | 1.9e-03 | 1.41 (1.14-1.75) | 1.7e-03 |
| **COVID-19-related distress** | | | | | | | | | | | | | | |
| (Intercept) | 1.92 (1.88-1.96) | 0.0e+00 | 2.81 (2.73-2.89) | 0.0e+00 | 3.88 (3.81-3.95) | 0.0e+00 | 4.35 (4.1-4.61) | 0.0e+00 | 0.22 (0.2-0.25) | 0.0e+00 | 0.11 (0.09-0.14) | 0.0e+00 | 0.02 (0.02-0.04) | 0.0e+00 |
| CV19RD | 0.97 (0.96-0.97) | 6.7e-16 | 0.93 (0.92-0.94) | 0.0e+00 | 0.93 (0.92-0.93) | 0.0e+00 | 0.72 (0.7-0.75) | 0.0e+00 | 1.32 (1.26-1.37) | 0.0e+00 | 2.03 (1.87-2.2) | 0.0e+00 | 3.23 (2.67-3.9) | 0.0e+00 |
| COVID-19 | 0.99 (0.98-1) | 1.5e-01 | 0.99 (0.97-1) | 5.7e-02 | 0.99 (0.98-1) | 5.9e-03 | 0.95 (0.93-0.98) | 3.0e-03 | 1.08 (1.03-1.13) | 1.9e-03 | 1.09 (0.99-1.21) | 8.2e-02 | 1.21 (0.99-1.49) | 6.5e-02 |
| Time | 1 (1-1) | 2.9e-03 | 1 (1-1) | 1.5e-02 | 1 (1-1) | 4.0e-04 | 0.99 (0.99-1) | 2.8e-06 | 1 (0.99-1) | 5.1e-01 | 1 (1-1.01) | 3.2e-01 | 1.03 (1.01-1.06) | 1.0e-03 |
| Sex (Male) | 1 (0.98-1.01) | 9.0e-01 | 1.02 (1-1.04) | 7.2e-02 | 1.01 (1-1.02) | 1.3e-01 | 1.03 (1-1.06) | 5.2e-02 | 0.96 (0.9-1.03) | 2.7e-01 | 0.96 (0.84-1.11) | 6.1e-01 | 1.01 (0.74-1.38) | 9.6e-01 |
| Age | 1 (1-1) | 9.3e-01 | 1 (1-1) | 1.9e-02 | 1 (1-1) | 4.9e-03 | 1 (1-1) | 9.2e-06 | 1 (1-1) | 4.0e-03 | 1 (1-1.01) | 2.9e-02 | 1 (1-1.01) | 7.1e-01 |
| Recruitment type* | 0.99 (0.98-1) | 1.6e-02 | 0.97 (0.95-0.98) | 2.2e-05 | 0.97 (0.97-0.98) | 2.7e-07 | 0.92 (0.9-0.95) | 1.6e-07 | 1.04 (0.99-1.09) | 1.5e-01 | 1.24 (1.12-1.37) | 2.7e-05 | 1.64 (1.32-2.03) | 7.5e-06 |

*Recruitment type: Self-recruitment

*Table S 2. Complete table of Relative Risk (RR) of better sleep quality and worst sleep quantity index using as predictors depression, anxiety, and COVID-19-related distress, adjusted for COVID-19, sex, age, time, and recruitment type.*

| | Sleep Quality | | | | | | | | Sleep Quantity Index | | | | | |
|---|---|---|---|---|---|---|---|---|---|---|---|---|---|---|
| | Score (2) | | Score (3) | | Score (4) | | Score (5) | | Score (1) | | Score (2) | | Score (3) | |
| | *RR (95% CI)* | *p-value* | *RR (95% CI)* | *p-value* | *RR (95% CI)* | *p-value* | *RR (95% CI)* | *p-value* | *RR (95% CI)* | *p-value* | *RR (95% CI)* | *p-value* | *RR (95% CI)* | *p-value* |
| **Depression** | | | | | | | | | | | | | | |
| (Intercept) | 2 (1.96-2.05) | 0.0e+00 | 3.08 (2.99-3.17) | 0.0e+00 | 4.08 (4.01-4.15) | 0.0e+00 | 4.85 (4.66-5.04) | 0.0e+00 | 0.21 (0.19-0.23) | 0.0e+00 | 0.08 (0.07-0.11) | 0.0e+00 | 0.01 (0.01-0.02) | 0.0e+00 |
| Depression | 0.91 (0.9-0.92) | 0.0e+00 | 0.79 (0.77-0.8) | 0.0e+00 | 0.66 (0.64-0.68) | 0.0e+00 | 0.32 (0.3-0.34) | 0.0e+00 | 1.74 (1.67-1.81) | 0.0e+00 | 4.04 (3.72-4.39) | 0.0e+00 | 10.46 (8.66-12.65) | 0.0e+00 |
| COVID-19 | 0.99 (0.98-1) | 1.3e-01 | 0.99 (0.98-1) | 1.2e-01 | 0.99 (0.98-1) | 5.9e-02 | 0.98 (0.96-1) | 2.8e-02 | 1.06 (1.01-1.11) | 1.2e-02 | 1.05 (0.95-1.15) | 3.2e-01 | 1.14 (0.94-1.39) | 1.8e-01 |
| Time | 1 (1-1) | 1.2e-03 | 1 (1-1) | 3.1e-02 | 1 (1-1) | 2.5e-04 | 1 (1-1) | 2.0e-03 | 1 (0.99-1) | 1.2e-01 | 1 (0.99-1.01) | 9.4e-01 | 1.02 (1-1.04) | 1.4e-02 |
| Sex (Male) | 1 (0.98-1.01) | 6.4e-01 | 1.01 (0.99-1.02) | 3.6e-01 | 1.01 (1-1.01) | 1.8e-01 | 1.04 (1.01-1.06) | 8.3e-04 | 0.96 (0.89-1.02) | 2.0e-01 | 0.96 (0.84-1.11) | 6.0e-01 | 1.05 (0.78-1.41) | 7.5e-01 |
| Age | 1 (1-1) | 7.1e-03 | 1 (1-1) | 2.8e-03 | 1 (1-1) | 1.5e-03 | 1 (1-1) | 8.7e-01 | 1 (1-1.01) | 1.2e-08 | 1.01 (1.01-1.01) | 1.3e-09 | 1.01 (1.01-1.02) | 2.7e-04 |
| Recruitment Type* | 1 (0.99-1.01) | 7.7e-01 | 0.99 (0.98-1) | 2.0e-01 | 1 (0.99-1.01) | 9.4e-01 | 0.99 (0.98-1.01) | 5.2e-01 | 0.98 (0.93-1.03) | 4.0e-01 | 1.05 (0.95-1.16) | 3.5e-01 | 1.1 (0.89-1.36) | 3.6e-01 |
| BMID (obese) | 0.99 (0.98-1.01) | 4.1e-01 | 0.98 (0.96-1) | 7.4e-02 | 0.99 (0.97-1) | 8.1e-02 | 0.99 (0.96-1.02) | 5.7e-01 | 1.17 (1.09-1.25) | 1.0e-05 | 1.31 (1.15-1.49) | 3.3e-05 | 1.58 (1.21-2.05) | 6.4e-04 |
| BMID (overweight) | 1 (0.99-1.01) | 4.0e-01 | 1 (0.99-1.01) | 9.8e-01 | 1 (0.99-1.01) | 9.4e-01 | 1 (0.98-1.01) | 6.3e-01 | 1.08 (1.02-1.14) | 4.0e-03 | 1.09 (0.98-1.21) | 1.3e-01 | 1.02 (0.8-1.29) | 8.8e-01 |
| Habitual Drinking** | 1 (0.99-1.01) | 7.3e-01 | 0.99 (0.98-1.01) | 4.8e-01 | 1 (0.99-1.01) | 5.7e-01 | 1.01 (0.99-1.03) | 2.9e-01 | 1.02 (0.97-1.07) | 5.1e-01 | 1.04 (0.93-1.15) | 4.8e-01 | 1.06 (0.85-1.32) | 6.2e-01 |
| Comorbidity (1) | 0.99 (0.98-1.01) | 2.8e-01 | 1 (0.98-1.01) | 9.4e-01 | 0.99 (0.98-1) | 1.8e-01 | 0.97 (0.95-1) | 2.3e-02 | 1.01 (0.96-1.07) | 6.5e-01 | 1.06 (0.95-1.18) | 3.2e-01 | 1.09 (0.86-1.39) | 4.7e-01 |
| Comorbidity (2) | 0.98 (0.95-1) | 2.3e-02 | 0.98 (0.95-1.01) | 1.2e-01 | 0.98 (0.96-1) | 2.8e-02 | 0.93 (0.89-0.98) | 4.4e-03 | 1 (0.91-1.1) | 9.6e-01 | 1.08 (0.91-1.28) | 4.0e-01 | 1.44 (1.02-2.04) | 4.1e-02 |
| Comorbidity (3+) | 0.99 (0.96-1.02) | 4.9e-01 | 1 (0.96-1.04) | 9.2e-01 | 1 (0.97-1.02) | 7.4e-01 | 0.95 (0.88-1.02) | 1.8e-01 | 1.01 (0.87-1.17) | 9.3e-01 | 1.08 (0.84-1.39) | 5.7e-01 | 1.64 (1-2.69) | 5.0e-02 |
| Psychiatric Diag | 1 (0.99-1.01) | 4.6e-01 | 0.99 (0.97-1) | 1.2e-01 | 0.98 (0.97-0.99) | 9.9e-04 | 0.93 (0.91-0.96) | 2.4e-06 | 1.04 (0.99-1.1) | 1.2e-01 | 1.08 (0.97-1.19) | 1.6e-01 | 1.34 (1.08-1.65) | 7.9e-03 |
| Relationship | 1 (0.99-1.01) | 5.4e-01 | 0.99 (0.97-1) | 3.9e-02 | 1 (0.99-1) | 3.4e-01 | 0.99 (0.97-1.01) | 2.0e-01 | 0.93 (0.89-0.98) | 7.2e-03 | 0.92 (0.83-1.01) | 7.9e-02 | 0.87 (0.71-1.07) | 1.9e-01 |
| Smoking | 0.99 (0.97-1) | 5.5e-02 | 0.98 (0.96-1) | 6.5e-02 | 0.98 (0.97-1) | 1.0e-02 | 0.96 (0.93-0.99) | 2.3e-02 | 1.11 (1.04-1.19) | 1.4e-03 | 1.2 (1.06-1.37) | 4.1e-03 | 1.39 (1.09-1.78) | 8.8e-03 |
| **Anxiety** | | | | | | | | | | | | | | |
| (Intercept) | 1.96 (1.91-2) | 0.0e+00 | 2.95 (2.86-3.04) | 0.0e+00 | 3.99 (3.92-4.06) | 0.0e+00 | 4.46 (4.22-4.72) | 0.0e+00 | 0.22 (0.2-0.25) | 0.0e+00 | 0.1 (0.08-0.13) | 0.0e+00 | 0.02 (0.01-0.03) | 0.0e+00 |
| Anxiety | 0.92 (0.91-0.93) | 0.0e+00 | 0.81 (0.79-0.83) | 0.0e+00 | 0.69 (0.67-0.72) | 0.0e+00 | 0.37 (0.34-0.39) | 0.0e+00 | 1.64 (1.56-1.72) | 0.0e+00 | 3.34 (3.06-3.66) | 0.0e+00 | 6.7 (5.52-8.13) | 0.0e+00 |
| COVID-19 | 0.99 (0.98-1) | 1.1e-01 | 0.99 (0.97-1) | 4.7e-02 | 0.99 (0.98-1) | 1.2e-02 | 0.97 (0.94-0.99) | 1.2e-02 | 1.07 (1.02-1.12) | 7.1e-03 | 1.08 (0.97-1.19) | 1.5e-01 | 1.19 (0.97-1.44) | 8.9e-02 |
| Time | 1 (1-1) | 2.7e-02 | 1 (1-1) | 3.4e-01 | 1 (1-1) | 7.3e-02 | 1 (1-1) | 4.3e-01 | 0.99 (0.99-1) | 2.3e-02 | 0.99 (0.99-1) | 2.5e-01 | 1.02 (1-1.04) | 7.0e-02 |
| Sex (Male) | 1 (0.99-1.02) | 9.7e-01 | 1.01 (1-1.03) | 1.5e-01 | 1.01 (1-1.02) | 6.0e-02 | 1.05 (1.02-1.08) | 1.9e-04 | 0.95 (0.88-1.01) | 1.2e-01 | 0.93 (0.81-1.07) | 3.0e-01 | 0.98 (0.72-1.33) | 9.0e-01 |
| Age | 1 (1-1) | 2.3e-02 | 1 (1-1) | 8.4e-02 | 1 (1-1) | 1.5e-02 | 1 (1-1) | 1.8e-01 | 1 (1-1.01) | 1.7e-07 | 1.01 (1.01-1.01) | 4.3e-08 | 1.01 (1-1.02) | 1.1e-03 |
| Recruitment Type* | 1 (0.99-1.01) | 4.7e-01 | 0.99 (0.97-1) | 3.8e-02 | 1 (0.99-1) | 3.3e-01 | 0.98 (0.95-1) | 4.9e-02 | 0.99 (0.94-1.04) | 6.8e-01 | 1.08 (0.98-1.2) | 1.4e-01 | 1.15 (0.93-1.43) | 2.0e-01 |
| BMID (obese) | 0.99 (0.97-1) | 9.0e-02 | 0.97 (0.94-0.99) | 5.2e-03 | 0.97 (0.96-0.99) | 2.1e-03 | 0.94 (0.9-0.98) | 7.4e-03 | 1.2 (1.12-1.28) | 2.5e-07 | 1.42 (1.25-1.62) | 1.2e-07 | 1.82 (1.4-2.38) | 8.5e-06 |
| BMID (overweight) | 1 (0.99-1.01) | 6.9e-01 | 1 (0.98-1.01) | 6.8e-01 | 1 (0.99-1.01) | 7.5e-01 | 0.99 (0.96-1.01) | 3.2e-01 | 1.09 (1.03-1.15) | 1.8e-03 | 1.11 (1-1.24) | 6.0e-02 | 1.07 (0.84-1.36) | 6.0e-01 |
| Habitual Drinking** | 1 (0.99-1.01) | 9.8e-01 | 0.99 (0.98-1.01) | 4.3e-01 | 1 (0.99-1.01) | 7.4e-01 | 1.01 (0.98-1.03) | 5.7e-01 | 1.01 (0.96-1.07) | 6.0e-01 | 1.03 (0.92-1.14) | 6.4e-01 | 1.02 (0.82-1.28) | 8.4e-01 |
| Comorbidity (1) | 0.99 (0.98-1) | 1.2e-01 | 1 (0.98-1.01) | 5.8e-01 | 0.99 (0.98-1) | 1.2e-01 | 0.97 (0.94-0.99) | 1.7e-02 | 1.02 (0.97-1.08) | 4.8e-01 | 1.08 (0.97-1.21) | 1.6e-01 | 1.13 (0.89-1.45) | 3.2e-01 |
| Comorbidity (2) | 0.97 (0.95-1) | 2.3e-02 | 0.97 (0.94-1) | 8.2e-02 | 0.97 (0.95-0.99) | 3.5e-03 | 0.91 (0.86-0.97) | 3.0e-03 | 1.01 (0.92-1.11) | 8.6e-01 | 1.14 (0.96-1.36) | 1.5e-01 | 1.57 (1.11-2.22) | 1.0e-02 |
| Comorbidity (3+) | 0.99 (0.96-1.02) | 3.5e-01 | 0.99 (0.95-1.03) | 6.6e-01 | 0.98 (0.95-1.01) | 2.1e-01 | 0.9 (0.81-1.01) | 6.5e-02 | 1.03 (0.89-1.2) | 6.6e-01 | 1.18 (0.91-1.53) | 2.1e-01 | 1.92 (1.16-3.17) | 1.1e-02 |
| Psychiatric Diag | 0.99 (0.98-1) | 1.1e-01 | 0.97 (0.96-0.99) | 1.2e-03 | 0.97 (0.96-0.98) | 1.0e-07 | 0.88 (0.85-0.92) | 4.4e-11 | 1.07 (1.02-1.13) | 5.3e-03 | 1.17 (1.05-1.3) | 3.0e-03 | 1.58 (1.26-1.99) | 6.4e-05 |
| Relationship | 1.01 (1-1.02) | 1.8e-01 | 1.01 (0.99-1.02) | 5.1e-01 | 1.01 (1-1.02) | 3.3e-02 | 1.03 (1-1.06) | 3.9e-02 | 0.9 (0.86-0.95) | 3.8e-05 | 0.82 (0.74-0.91) | 1.1e-04 | 0.72 (0.59-0.89) | 2.4e-03 |
| Smoking | 0.98 (0.97-1) | 4.0e-02 | 0.98 (0.96-1) | 4.5e-02 | 0.97 (0.96-0.99) | 8.0e-04 | 0.96 (0.92-1) | 2.9e-02 | 1.13 (1.06-1.21) | 3.1e-04 | 1.26 (1.11-1.44) | 4.1e-04 | 1.49 (1.16-1.91) | 2.0e-03 |
| **COVID-19-related distress** | | | | | | | | | | | | | | |
| (Intercept) | 1.93 (1.88-1.97) | 0.0e+00 | 2.87 (2.78-2.97) | 0.0e+00 | 3.91 (3.83-3.99) | 0.0e+00 | 4.43 (4.14-4.73) | 0.0e+00 | 0.22 (0.2-0.25) | 0.0e+00 | 0.11 (0.09-0.14) | 0.0e+00 | 0.02 (0.02-0.04) | 0.0e+00 |
| CV19RD | 0.97 (0.96-0.98) | 2.7e-12 | 0.94 (0.93-0.95) | 0.0e+00 | 0.93 (0.93-0.94) | 0.0e+00 | 0.75 (0.73-0.78) | 0.0e+00 | 1.28 (1.23-1.33) | 0.0e+00 | 1.89 (1.74-2.05) | 0.0e+00 | 2.71 (2.24-3.28) | 0.0e+00 |
| COVID-19 | 0.99 (0.98-1) | 1.6e-01 | 0.99 (0.97-1) | 8.8e-02 | 0.99 (0.98-1) | 8.6e-03 | 0.96 (0.93-0.99) | 6.3e-03 | 1.07 (1.02-1.12) | 5.2e-03 | 1.08 (0.98-1.19) | 1.4e-01 | 1.18 (0.96-1.45) | 1.1e-01 |
| Time | 1 (1-1) | 3.7e-03 | 1 (1-1) | 1.7e-02 | 1 (1-1) | 1.1e-03 | 0.99 (0.99-1) | 2.3e-05 | 1 (0.99-1) | 4.9e-01 | 1 (0.99-1.01) | 4.5e-01 | 1.03 (1.01-1.05) | 2.7e-03 |
| Sex (Male) | 1 (0.98-1.01) | 8.6e-01 | 1.01 (0.99-1.03) | 1.8e-01 | 1.01 (1-1.02) | 2.7e-01 | 1.03 (1-1.06) | 4.5e-02 | 0.97 (0.9-1.03) | 3.2e-01 | 0.97 (0.84-1.12) | 6.7e-01 | 1.02 (0.75-1.4) | 8.9e-01 |
| Age | 1 (1-1) | 9.5e-01 | 1 (1-1) | 7.2e-02 | 1 (1-1) | 3.5e-03 | 1 (1-1) | 3.8e-06 | 1 (1-1) | 4.3e-03 | 1 (1-1.01) | 5.9e-02 | 1 (0.99-1.01) | 9.9e-01 |
| Recruitment Type* | 0.99 (0.98-1) | 2.4e-01 | 0.98 (0.96-0.99) | 6.2e-03 | 0.99 (0.98-1) | 9.7e-03 | 0.96 (0.93-0.99) | 3.4e-03 | 1 (0.95-1.05) | 8.5e-01 | 1.12 (1.01-1.24) | 3.7e-02 | 1.26 (1.01-1.56) | 3.9e-02 |
| BMID (obese) | 0.98 (0.97-1) | 8.0e-02 | 0.96 (0.94-0.99) | 6.8e-03 | 0.97 (0.95-0.98) | 4.1e-04 | 0.92 (0.87-0.97) | 2.9e-03 | 1.2 (1.12-1.28) | 3.2e-07 | 1.44 (1.26-1.64) | 8.7e-08 | 1.86 (1.41-2.45) | 1.2e-05 |
| BMID (overweight) | 1 (0.99-1.01) | 5.0e-01 | 1 (0.98-1.01) | 9.0e-01 | 1 (0.99-1.01) | 9.0e-01 | 0.98 (0.95-1.01) | 1.9e-01 | 1.08 (1.03-1.14) | 3.4e-03 | 1.09 (0.98-1.22) | 1.2e-01 | 1.02 (0.8-1.3) | 8.9e-01 |
| Habitual Drinking** | 1 (0.99-1.01) | 9.4e-01 | 1 (0.98-1.01) | 7.0e-01 | 1 (0.99-1.01) | 9.7e-01 | 1.01 (0.98-1.04) | 5.4e-01 | 1.01 (0.96-1.06) | 7.9e-01 | 1.01 (0.9-1.13) | 8.7e-01 | 0.97 (0.77-1.22) | 8.1e-01 |
| Comorbidity (1) | 0.99 (0.98-1) | 1.8e-01 | 0.99 (0.97-1.01) | 3.1e-01 | 0.99 (0.98-1) | 2.0e-02 | 0.96 (0.93-0.99) | 1.7e-02 | 1.02 (0.97-1.08) | 4.8e-01 | 1.11 (0.99-1.24) | 7.9e-02 | 1.2 (0.94-1.54) | 1.5e-01 |
| Comorbidity (2) | 0.97 (0.95-1) | 1.9e-02 | 0.97 (0.94-1) | 4.7e-02 | 0.97 (0.94-0.99) | 3.6e-03 | 0.89 (0.83-0.95) | 1.2e-03 | 1 (0.91-1.1) | 9.7e-01 | 1.14 (0.95-1.37) | 1.5e-01 | 1.64 (1.13-2.37) | 9.3e-03 |
| Comorbidity (3+) | 0.98 (0.95-1.02) | 3.0e-01 | 0.98 (0.94-1.02) | 3.5e-01 | 0.96 (0.93-1) | 3.6e-02 | 0.86 (0.76-0.98) | 1.9e-02 | 1.05 (0.9-1.22) | 5.4e-01 | 1.27 (0.97-1.66) | 7.9e-02 | 2.27 (1.35-3.83) | 2.0e-03 |
| Psychiatric Diag | 0.98 (0.97-0.99) | 7.2e-04 | 0.95 (0.93-0.97) | 3.4e-09 | 0.94 (0.93-0.96) | 0.0e+00 | 0.81 (0.77-0.84) | 0.0e+00 | 1.12 (1.06-1.18) | 9.6e-06 | 1.35 (1.22-1.49) | 7.3e-09 | 2.09 (1.68-2.6) | 4.2e-11 |
| Relationship | 1.01 (1-1.02) | 1.6e-01 | 1.01 (0.99-1.02) | 3.4e-01 | 1.01 (1-1.02) | 3.9e-02 | 1.04 (1.01-1.08) | 1.9e-02 | 0.9 (0.86-0.95) | 5.0e-05 | 0.82 (0.74-0.9) | 8.0e-05 | 0.7 (0.57-0.87) | 1.2e-03 |
| Smoking | 0.98 (0.97-1) | 5.0e-02 | 0.97 (0.95-1) | 3.3e-02 | 0.97 (0.95-0.99) | 3.0e-04 | 0.92 (0.88-0.97) | 8.2e-04 | 1.13 (1.05-1.2) | 4.2e-04 | 1.27 (1.11-1.44) | 4.0e-04 | 1.56 (1.2-2.01) | 7.7e-04 |

*Recruitment type: Self-recuitment

** For women, 4 or more drinks consumed on one occasion (one occasion = 2-3 hours). For men, 5 or more drinks consumed on one occasion.

*Table S 3. Complete table of Relative Risk (RR) of better sleep quality and worst sleep quantity index using as predictors depression, anxiety, and COVID-19-related distress, adjusted for COVID-19, sex, age, time, recruitment type, body mass index (BMID), habitual drinking, comorbidities, previous psychiatric diagnosis relationship, and smoking.*

| | Depression | | Anxiety | | COVID-19-related distress | | Stress | |
|---|---|---|---|---|---|---|---|---|
| | *RR (95% CI)* | *p-value* | *RR (95% CI)* | *p-value* | *RR (95% CI)* | *p-value* | *RR (95% CI)* | *p-value* |
| **Model Sleep Quant Scale** | | | | | | | | |
| (Intercept) | 6.44 (6.01-6.9) | 0.0e+00 | 6.16 (5.69-6.66) | 0.00e+00 | 3.78 (3.55-4.03) | 0.00e+00 | 7.93 (7.8-8.06) | 0.00e+00 |
| SQS (5< hours) | 3.23 (3.03-3.44) | 0.0e+00 | 3.04 (2.81-3.3) | 0.00e+00 | 1.81 (1.69-1.93) | 0.00e+00 | 1.09 (1.06-1.12) | 6.76e-11 |
| SQS (5 or 6 hours) | 1.77 (1.73-1.82) | 0.0e+00 | 1.73 (1.68-1.79) | 0.00e+00 | 1.3 (1.26-1.33) | 0.00e+00 | 1.04 (1.03-1.04) | 0.00e+00 |
| SQS (10 or 11 hours) | 2.34 (2.16-2.54) | 0.0e+00 | 1.82 (1.64-2.03) | 0.00e+00 | 1.4 (1.28-1.54) | 2.60e-13 | 1.03 (1-1.06) | 6.32e-02 |
| SQS (>11 hours) | 3.17 (2.83-3.56) | 0.0e+00 | 2.38 (2-2.83) | 0.00e+00 | 1.94 (1.65-2.26) | 1.11e-16 | 1.1 (1.04-1.17) | 1.81e-03 |
| COVID-19 | 1 (0.97-1.04) | 8.5e-01 | 0.97 (0.93-1.01) | 9.88e-02 | 0.92 (0.89-0.95) | 1.18e-06 | 0.98 (0.97-0.99) | 1.94e-04 |
| Time | 0.98 (0.98-0.99) | 0.0e+00 | 0.99 (0.99-1) | 4.74e-06 | 0.97 (0.97-0.98) | 0.00e+00 | 1 (1-1) | 3.41e-01 |
| Sex (Male) | 0.75 (0.71-0.79) | 0.0e+00 | 0.69 (0.65-0.73) | 0.00e+00 | 0.69 (0.66-0.72) | 0.00e+00 | 0.91 (0.9-0.93) | 0.00e+00 |
| Age | 0.99 (0.99-0.99) | 0.0e+00 | 0.98 (0.98-0.98) | 0.00e+00 | 1 (1-1) | 2.93e-05 | 1 (1-1) | 0.00e+00 |
| Recruitment Type* | 1.34 (1.3-1.39) | 0.0e+00 | 1.36 (1.3-1.41) | 0.00e+00 | 1.24 (1.2-1.28) | 0.00e+00 | 1.03 (1.03-1.04) | 1.38e-14 |
| **Model Sleep Quant Index** | | | | | | | | |
| (Intercept) | 6.53 (6.1-6.99) | 0.0e+00 | 6.2 (5.73-6.71) | 0.00e+00 | 3.79 (3.56-4.04) | 0.00e+00 | 7.93 (7.8-8.07) | 0.00e+00 |
| SQI (6 or 10 hours) | 1.62 (1.58-1.66) | 0.0e+00 | 1.58 (1.53-1.63) | 0.00e+00 | 1.24 (1.21-1.27) | 0.00e+00 | 1.03 (1.02-1.04) | 1.01e-14 |
| SQI (5 or 11 hours) | 2.35 (2.26-2.44) | 0.0e+00 | 2.24 (2.14-2.34) | 0.00e+00 | 1.5 (1.44-1.55) | 0.00e+00 | 1.06 (1.05-1.07) | 0.00e+00 |
| SQI (5< or >11 hours) | 3.23 (3.04-3.43) | 0.0e+00 | 3.01 (2.78-3.25) | 0.00e+00 | 1.81 (1.7-1.93) | 0.00e+00 | 1.09 (1.06-1.12) | 9.12e-12 |
| COVID-19 | 1 (0.97-1.04) | 8.6e-01 | 0.97 (0.93-1.01) | 1.02e-01 | 0.92 (0.89-0.95) | 1.20e-06 | 0.98 (0.97-0.99) | 2.03e-04 |
| Time | 0.98 (0.98-0.99) | 0.0e+00 | 0.99 (0.99-1) | 2.97e-06 | 0.97 (0.97-0.98) | 0.00e+00 | 1 (1-1) | 3.35e-01 |
| Sex (Male) | 0.75 (0.71-0.79) | 0.0e+00 | 0.69 (0.65-0.74) | 0.00e+00 | 0.69 (0.66-0.72) | 0.00e+00 | 0.91 (0.9-0.93) | 0.00e+00 |
| Age | 0.99 (0.99-0.99) | 0.0e+00 | 0.98 (0.98-0.98) | 0.00e+00 | 1 (1-1) | 4.13e-05 | 1 (1-1) | 0.00e+00 |
| Recruitment Type* | 1.34 (1.29-1.38) | 0.0e+00 | 1.35 (1.29-1.4) | 0.00e+00 | 1.24 (1.2-1.28) | 0.00e+00 | 1.03 (1.02-1.04) | 3.36e-14 |

*Model:* Adjusted COVID-19, Sex, Time & Recruitment type
*Recruitment type: Self-recruitment

*Table S 4 Table of Relative Risk (RR) of depression, anxiety, Covid-19 related distress, and stress using as predictors sleep quantity scale and sleep quantity index.*

| | Sleep Quantity Scale* | | | | | | | | Sleep Quantity Index* | | | | | |
|---|---|---|---|---|---|---|---|---|---|---|---|---|---|---|
| | **Score (5< hours)** | | **Score (5 or 6 hours)** | | **Score (10 or 11 hours)** | | **Score (>11 hours)** | | Score (6 or 10 hours) | | Score (5 or 11 hours) | | Score (5< or >11 hours) | |
| | *RR (95% CI)* | *p-value* | *RR (95% CI)* | *p-value* | *RR (95% CI)* | *p-value* | *RR (95% CI)* | *p-value* | *RR (95% CI)* | *p-value* | *RR (95% CI)* | *p-value* | *RR (95% CI)* | *p-value* |
| **Depression** | | | | | | | | | | | | | | |
| (Intercept) | 0 (0-0.01) | 0.0e+00 | 0.45 (0.41-0.49) | 0.0e+00 | 0.03 (0.02-0.06) | 0.0e+00 | 0 (0-0) | 3.6e-13 | 0.21 (0.19-0.23) | 0.0e+00 | 0.08 (0.07-0.1) | 0.0e+00 | 0.01 (0.01-0.02) | 0.0e+00 |
| Depression | 12.78 (10.6-15.4) | 0.0e+00 | 1.9 (1.84-1.97) | 0.0e+00 | 7.05 (5.74-8.64) | 0.0e+00 | 49.35 (11.09-219.48) | 3.0e-07 | 1.81 (1.74-1.88) | 0.0e+00 | 4.36 (4.03-4.71) | 0.0e+00 | 12.94 (10.8-15.51) | 0.0e+00 |
| COVID-19 | 1.14 (0.93-1.39) | 2.1e-01 | 1.06 (1.02-1.11) | 6.9e-03 | 0.94 (0.73-1.2) | 6.1e-01 | 3.14 (0.95-10.46) | 6.2e-02 | 1.07 (1.02-1.12) | 6.5e-03 | 1.05 (0.96-1.16) | 2.9e-01 | 1.14 (0.94-1.39) | 1.7e-01 |
| Time | 1.03 (1.01-1.05) | 7.3e-03 | 1 (0.99-1) | 1.8e-01 | 1 (0.97-1.03) | 9.0e-01 | 1.01 (0.8-1.27) | 9.3e-01 | 1 (0.99-1) | 1.1e-01 | 1 (0.99-1.01) | 9.7e-01 | 1.03 (1.01-1.05) | 6.7e-03 |
| Sex (Male) | 1.09 (0.8-1.47) | 5.9e-01 | 0.97 (0.92-1.04) | 4.2e-01 | 0.53 (0.35-0.81) | 3.5e-03 | 0.5 (0.07-3.82) | 5.1e-01 | 0.96 (0.9-1.03) | 2.4e-01 | 0.98 (0.85-1.12) | 7.3e-01 | 1.07 (0.8-1.43) | 6.6e-01 |
| Age | 1.02 (1.01-1.02) | 5.2e-08 | 1.01 (1-1.01) | 1.2e-14 | 1 (0.99-1) | 2.9e-01 | 0.99 (0.95-1.03) | 6.0e-01 | 1 (1-1.01) | 1.6e-10 | 1.01 (1.01-1.01) | 4.8e-13 | 1.01 (1.01-1.02) | 3.2e-07 |
| Recruitment Type** | 1.25 (1.01-1.55) | 4.1e-02 | 1.01 (0.97-1.06) | 5.7e-01 | 1.5 (1.13-1.99) | 5.1e-03 | 2.04 (0.67-6.18) | 2.1e-01 | 1 (0.96-1.05) | 8.9e-01 | 1.1 (1-1.21) | 5.1e-02 | 1.26 (1.03-1.56) | 2.7e-02 |
| **Anxiety** | | | | | | | | | | | | | | |
| (Intercept) | 0.01 (0-0.01) | 0.0e+00 | 0.48 (0.44-0.53) | 0.0e+00 | 0.05 (0.03-0.1) | 0.0e+00 | 0 (0-0.01) | 2.4e-09 | 0.22 (0.2-0.24) | 0.0e+00 | 0.1 (0.08-0.12) | 0.0e+00 | 0.02 (0.01-0.03) | 0.0e+00 |
| Anxiety | 8.98 (7.45-10.83) | 0.0e+00 | 1.8 (1.73-1.87) | 0.0e+00 | 3.55 (2.77-4.55) | 0.0e+00 | 12.94 (4.2-39.92) | 8.4e-06 | 1.7 (1.63-1.78) | 0.0e+00 | 3.7 (3.4-4.03) | 0.0e+00 | 8.79 (7.33-10.53) | 0.0e+00 |
| COVID-19 | 1.19 (0.97-1.46) | 9.9e-02 | 1.07 (1.02-1.12) | 2.5e-03 | 0.96 (0.74-1.25) | 7.7e-01 | 3.4 (0.99-11.75) | 5.2e-02 | 1.07 (1.02-1.13) | 3.4e-03 | 1.08 (0.98-1.19) | 1.2e-01 | 1.2 (0.98-1.46) | 7.6e-02 |
| Time | 1.02 (1-1.04) | 6.3e-02 | 1 (0.99-1) | 9.5e-03 | 0.99 (0.96-1.02) | 5.1e-01 | 0.99 (0.78-1.24) | 9.1e-01 | 0.99 (0.99-1) | 1.6e-02 | 0.99 (0.99-1) | 2.6e-01 | 1.02 (1-1.04) | 6.6e-02 |
| Sex (Male) | 1 (0.74-1.37) | 9.8e-01 | 0.96 (0.9-1.02) | 1.8e-01 | 0.48 (0.31-0.74) | 8.6e-04 | 0.44 (0.06-3.25) | 4.2e-01 | 0.95 (0.88-1.01) | 1.1e-01 | 0.93 (0.81-1.07) | 3.3e-01 | 0.98 (0.73-1.33) | 9.1e-01 |
| Age | 1.02 (1.01-1.02) | 2.7e-07 | 1.01 (1-1.01) | 5.7e-13 | 0.99 (0.98-1) | 6.0e-02 | 0.99 (0.94-1.03) | 5.1e-01 | 1 (1-1.01) | 4.3e-09 | 1.01 (1.01-1.01) | 1.7e-11 | 1.01 (1.01-1.02) | 2.3e-06 |
| Recruitment Type* | 1.39 (1.11-1.73) | 3.7e-03 | 1.04 (0.99-1.09) | 1.0e-01 | 1.7 (1.27-2.27) | 3.1e-04 | 2.43 (0.78-7.55) | 1.3e-01 | 1.02 (0.98-1.08) | 3.2e-01 | 1.17 (1.06-1.29) | 1.9e-03 | 1.41 (1.14-1.75) | 1.7e-03 |
| **COVID-19-related distress** | | | | | | | | | | | | | | |
| (Intercept) | 0.01 (0-0.01) | 0.0e+00 | 0.49 (0.45-0.54) | 0.0e+00 | 0.06 (0.03-0.11) | 0.0e+00 | 0 (0-0.01) | 8.9e-09 | 0.22 (0.2-0.25) | 0.0e+00 | 0.11 (0.09-0.14) | 0.0e+00 | 0.02 (0.02-0.04) | 0.0e+00 |
| CV19RD | 3.18 (2.62-3.87) | 0.0e+00 | 1.07 (1.03-1.12) | 1.4e-03 | 1.97 (1.56-2.5) | 1.6e-08 | 8.58 (1.87-39.23) | 5.6e-03 | 1.32 (1.26-1.37) | 0.0e+00 | 2.03 (1.87-2.2) | 0.0e+00 | 3.23 (2.67-3.9) | 0.0e+00 |
| COVID-19 | 1.2 (0.97-1.49) | 8.9e-02 | 1 (1-1) | 8.5e-01 | 0.97 (0.74-1.25) | 7.9e-01 | 3.4 (0.98-11.8) | 5.4e-02 | 1.08 (1.03-1.13) | 1.9e-03 | 1.09 (0.99-1.21) | 8.2e-02 | 1.21 (0.99-1.49) | 6.5e-02 |
| Time | 1.04 (1.01-1.06) | 1.1e-03 | 0.98 (0.92-1.04) | 4.6e-01 | 1 (0.97-1.03) | 8.6e-01 | 1.01 (0.8-1.27) | 9.5e-01 | 1 (0.99-1) | 5.1e-01 | 1 (1-1.01) | 3.2e-01 | 1.03 (1.01-1.06) | 1.0e-03 |
| Sex (Male) | 1.03 (0.75-1.41) | 8.6e-01 | 1 (1-1) | 4.6e-04 | 0.49 (0.32-0.76) | 1.3e-03 | 0.44 (0.06-3.47) | 4.4e-01 | 0.96 (0.9-1.03) | 2.7e-01 | 0.96 (0.84-1.11) | 6.1e-01 | 1.01 (0.74-1.38) | 9.6e-01 |
| Age | 1 (1-1.01) | 3.8e-01 | 1.05 (1.01-1.1) | 2.5e-02 | 0.98 (0.97-0.99) | 6.6e-04 | 0.97 (0.93-1.01) | 1.4e-01 | 1 (1-1) | 4.0e-03 | 1 (1-1.01) | 2.9e-02 | 1 (1-1.01) | 7.1e-01 |
| Recruitment Type** | 1.62 (1.29-2.02) | 2.3e-05 | 1.07 (1.03-1.12) | 1.4e-03 | 1.79 (1.34-2.39) | 8.5e-05 | 2.78 (0.93-8.35) | 6.8e-02 | 1.04 (0.99-1.09) | 1.5e-01 | 1.24 (1.12-1.37) | 2.7e-05 | 1.64 (1.32-2.03) | 7.5e-06 |

*Sleep quantity always compare with normal sleep from 7 to 9 hours
**Recruitment Type: Self-recruitment

*Table S 5 Table of Relative Risk (RR) of sleep quantity scale and sleep quantity index using as predictors depression, anxiety, and COVID-19 related distress, adjusted for COVID-19, sex, age, time, and recruitment type.*

| | Depression | | Anxiety | | COVID-19-related distress | | Stress | |
|---|---|---|---|---|---|---|---|---|
| | *RR (95% CI)* | *p-value* | *RR (95% CI)* | *p-value* | *RR (95% CI)* | *p-value* | *RR (95% CI)* | *p-value* |
| **Model Sleep Quality** | | | | | | | | |
| (Intercept) | 19.78 (18.52-21.12) | 0.00e+00 | 17.92 (16.54-19.43) | 0.00e+00 | 6.83 (6.36-7.34) | 0.00e+00 | 8.75 (8.55-8.95) | 0.00e+00 |
| Sleep Quality (2) | 0.66 (0.64-0.68) | 0.00e+00 | 0.66 (0.63-0.7) | 0.00e+00 | 0.77 (0.74-0.81) | 0.00e+00 | 0.96 (0.94-0.97) | 1.85e-07 |
| Sleep Quality (3) | 0.47 (0.45-0.49) | 0.00e+00 | 0.48 (0.45-0.51) | 0.00e+00 | 0.66 (0.63-0.69) | 0.00e+00 | 0.92 (0.9-0.93) | 0.00e+00 |
| Sleep Quality (4) | 0.25 (0.24-0.26) | 0.00e+00 | 0.27 (0.26-0.29) | 0.00e+00 | 0.5 (0.48-0.53) | 0.00e+00 | 0.89 (0.88-0.91) | 0.00e+00 |
| Sleep Quality (5) | 0.13 (0.12-0.14) | 0.00e+00 | 0.15 (0.14-0.16) | 0.00e+00 | 0.38 (0.35-0.4) | 0.00e+00 | 0.88 (0.86-0.9) | 0.00e+00 |
| **Model Sleep Quality Female** | | | | | | | | |
| (Intercept) | 18.85 (17.56-20.23) | 0.00e+00 | 17.08 (15.66-18.63) | 0.00e+00 | 6.73 (6.23-7.27) | 0.00e+00 | 8.55 (8.34-8.75) | 0.00e+00 |
| Sleep Quality (2) | 0.67 (0.64-0.69) | 0.00e+00 | 0.67 (0.64-0.71) | 0.00e+00 | 0.78 (0.75-0.82) | 0.00e+00 | 0.96 (0.95-0.98) | 3.81e-06 |
| Sleep Quality (3) | 0.48 (0.46-0.5) | 0.00e+00 | 0.49 (0.47-0.52) | 0.00e+00 | 0.67 (0.64-0.71) | 0.00e+00 | 0.92 (0.91-0.94) | 0.00e+00 |
| Sleep Quality (4) | 0.26 (0.25-0.27) | 0.00e+00 | 0.29 (0.27-0.3) | 0.00e+00 | 0.52 (0.49-0.54) | 0.00e+00 | 0.9 (0.88-0.91) | 0.00e+00 |
| Sleep Quality (5) | 0.14 (0.13-0.15) | 0.00e+00 | 0.16 (0.14-0.17) | 0.00e+00 | 0.39 (0.37-0.42) | 0.00e+00 | 0.89 (0.87-0.9) | 0.00e+00 |
| **Model Sleep Quality Male** | | | | | | | | |
| (Intercept) | 21.88 (18.24-26.26) | 0.00e+00 | 18.96 (15.21-23.63) | 0.00e+00 | 5.71 (4.68-6.96) | 0.00e+00 | 9.03 (8.51-9.57) | 0.00e+00 |
| Sleep Quality (2) | 0.61 (0.53-0.7) | 2.21e-13 | 0.59 (0.5-0.69) | 3.40e-11 | 0.7 (0.61-0.81) | 9.39e-07 | 0.94 (0.9-0.98) | 8.64e-03 |
| Sleep Quality (3) | 0.39 (0.34-0.45) | 0.00e+00 | 0.37 (0.31-0.44) | 0.00e+00 | 0.56 (0.49-0.65) | 3.92e-14 | 0.89 (0.85-0.93) | 3.80e-07 |
| Sleep Quality (4) | 0.2 (0.17-0.23) | 0.00e+00 | 0.2 (0.16-0.24) | 0.00e+00 | 0.39 (0.33-0.45) | 0.00e+00 | 0.86 (0.82-0.9) | 3.91e-10 |
| Sleep Quality (5) | 0.09 (0.08-0.12) | 0.00e+00 | 0.09 (0.07-0.11) | 0.00e+00 | 0.27 (0.23-0.32) | 0.00e+00 | 0.83 (0.79-0.88) | 1.02e-11 |
| **Model Sleep Quantity** | | | | | | | | |
| (Intercept) | 6.53 (6.1-6.99) | 0.0e+00 | 6.2 (5.73-6.71) | 0.00e+00 | 3.79 (3.56-4.04) | 0.00e+00 | 7.93 (7.8-8.07) | 0.00e+00 |
| SQI (6 or 10 hours) | 1.62 (1.58-1.66) | 0.0e+00 | 1.58 (1.53-1.63) | 0.00e+00 | 1.24 (1.21-1.27) | 0.00e+00 | 1.03 (1.02-1.04) | 1.01e-14 |
| SQI (5 or 11 hours) | 2.35 (2.26-2.44) | 0.0e+00 | 2.24 (2.14-2.34) | 0.00e+00 | 1.5 (1.44-1.55) | 0.00e+00 | 1.06 (1.05-1.07) | 0.00e+00 |
| SQI (5< or >11 hours) | 3.23 (3.04-3.43) | 0.0e+00 | 3.01 (2.78-3.25) | 0.00e+00 | 1.81 (1.7-1.93) | 0.00e+00 | 1.09 (1.06-1.12) | 9.12e-12 |
| **Model Sleep Quantity Female** | | | | | | | | |
| (Intercept) | 6.33 (5.88-6.82) | 0.00e+00 | 6.07 (5.58-6.61) | 0.00e+00 | 3.8 (3.55-4.07) | 0.00e+00 | 7.79 (7.65-7.93) | 0.00e+00 |
| SQI (6 or 10 hours) | 1.62 (1.57-1.66) | 0.00e+00 | 1.57 (1.52-1.62) | 0.00e+00 | 1.23 (1.2-1.27) | 0.00e+00 | 1.03 (1.02-1.03) | 1.20e-11 |
| SQI (5 or 11 hours) | 2.31 (2.22-2.4) | 0.00e+00 | 2.19 (2.09-2.3) | 0.00e+00 | 1.48 (1.43-1.54) | 0.00e+00 | 1.06 (1.05-1.07) | 0.00e+00 |
| SQI (5< or >11 hours) | 3.04 (2.87-3.22) | 0.00e+00 | 2.83 (2.62-3.05) | 0.00e+00 | 1.76 (1.65-1.88) | 0.00e+00 | 1.08 (1.05-1.11) | 2.31e-09 |
| **Model Sleep Quantity Male** | | | | | | | | |
| (Intercept) | 6.11 (5.15-7.25) | 0.00e+00 | 5.19 (4.26-6.32) | 0.00e+00 | 2.63 (2.23-3.12) | 0.00e+00 | 7.87 (7.55-8.22) | 0.00e+00 |
| SQI (6 or 10 hours) | 1.64 (1.51-1.77) | 0.00e+00 | 1.63 (1.47-1.79) | 0.00e+00 | 1.26 (1.16-1.36) | 5.58e-09 | 1.04 (1.02-1.06) | 2.58e-04 |
| SQI (5 or 11 hours) | 2.65 (2.36-2.97) | 0.00e+00 | 2.55 (2.21-2.94) | 0.00e+00 | 1.63 (1.45-1.84) | 5.55e-16 | 1.09 (1.06-1.13) | 2.16e-09 |
| SQI (5< or >11 hours) | 4.93 (3.99-6.09) | 0.00e+00 | 4.85 (3.71-6.36) | 0.00e+00 | 2.29 (1.84-2.84) | 1.13e-13 | 1.15 (1.06-1.25) | 7.65e-04 |

*Table S 6 Table of Relative Risk (RR) of depression, anxiety, Covid-19 related distress, and stress using as predictors sleep quality and sleep quantity index. Stratified by sex.*

| | Sleep Quality | | | | | | | | Sleep Quantity Index | | | | | |
|---|---|---|---|---|---|---|---|---|---|---|---|---|---|---|
| | Score (Poor) | | Score (Medium) | | Score (Good) | | Score (Very Good) | | Score (6 or 10 hours) | | Score (5 or 11 hours) | | Score (5< or >11 hours) | |
| | *RR (95% CI)* | *p-value* | *RR (95% CI)* | *p-value* | *RR (95% CI)* | *p-value* | *RR (95% CI)* | *p-value* | *RR (95% CI)* | *p-value* | *RR (95% CI)* | *p-value* | *RR (95% CI)* | *p-value* |
| **Depression** | | | | | | | | | | | | | | |
| (Intercept) | 2.00 (1.95-2.04) | 0.0e+00 | 3.02 (2.94-3.1) | 0.0e+00 | 4.04 (3.98-4.1) | 0.0e+00 | 4.76 (4.59-4.93) | 0.0e+00 | 0.21 (0.19-0.23) | 0.0e+00 | 0.08 (0.07-0.1) | 0.0e+00 | 0.01 (0.01-0.02) | 0.0e+00 |
| Depression | 0.91 (0.9-0.92) | 0.0e+00 | 0.78 (0.77-0.8) | 0.0e+00 | 0.66 (0.64-0.68) | 0.0e+00 | 0.31 (0.29-0.32) | 0.0e+00 | 1.81 (1.74-1.88) | 0.0e+00 | 4.36 (4.03-4.71) | 0.0e+00 | 12.94 (10.8-15.51) | 0.0e+00 |
| **Depression Female** | | | | | | | | | | | | | | |
| (Intercept) | 1.99 (1.95-2.04) | 0.0e+00 | 3.01 (2.93-3.1) | 0.0e+00 | 4.06 (4-4.13) | 0.0e+00 | 4.8 (4.6-5.01) | 0.0e+00 | 0.2 (0.18-0.22) | 0.0e+00 | 0.08 (0.07-0.1) | 0.0e+00 | 0.01 (0.01-0.02) | 0.0e+00 |
| Depression | 0.91 (0.9-0.92) | 0.0e+00 | 0.79 (0.77-0.8) | 0.0e+00 | 0.66 (0.64-0.68) | 0.0e+00 | 0.31 (0.29-0.33) | 0.0e+00 | 1.81 (1.73-1.89) | 0.0e+00 | 4.28 (3.94-4.65) | 0.0e+00 | 11.8 (9.79-14.24) | 0.0e+00 |
| **Depression Male** | | | | | | | | | | | | | | |
| (Intercept) | 1.99 (1.88-2.11) | 0.0e+00 | 3.05 (2.88-3.23) | 0.0e+00 | 3.98 (3.87-4.09) | 0.0e+00 | 4.73 (4.42-5.07) | 0.0e+00 | 0.23 (0.18-0.29) | 0.0e+00 | 0.1 (0.06-0.16) | 0.0e+00 | 0 (0-0.02) | 0.0e+00 |
| Depression | 0.89 (0.87-0.92) | 9.9e-12 | 0.76 (0.71-0.81) | 5.6e-16 | 0.63 (0.57-0.7) | 0.0e+00 | 0.3 (0.26-0.34) | 0.0e+00 | 1.82 (1.6-2.06) | 0.0e+00 | 4.88 (3.9-6.1) | 0.0e+00 | 22.57 (13.27-38.4) | 0.0e+00 |
| **Anxiety** | | | | | | | | | | | | | | |
| (Intercept) | 1.96 (1.92-2) | 0.0e+00 | 2.91 (2.83-2.99) | 0.0e+00 | 3.99 (3.92-4.05) | 0.0e+00 | 4.47 (4.26-4.69) | 0.0e+00 | 0.22 (0.2-0.24) | 0.0e+00 | 0.1 (0.08-0.12) | 0.0e+00 | 0.02 (0.01-0.03) | 0.0e+00 |
| Anxiety | 0.92 (0.91-0.93) | 0.0e+00 | 0.8 (0.78-0.82) | 0.0e+00 | 0.68 (0.66-0.71) | 0.0e+00 | 0.34 (0.32-0.36) | 0.0e+00 | 1.7 (1.63-1.78) | 0.0e+00 | 3.7 (3.4-4.03) | 0.0e+00 | 8.79 (7.33-10.53) | 0.0e+00 |
| **Anxiety Female** | | | | | | | | | | | | | | |
| (Intercept) | 1.96 (1.91-2) | 0.0e+00 | 2.91 (2.83-3) | 0.0e+00 | 4.02 (3.94-4.09) | 0.0e+00 | 4.54 (4.29-4.8) | 0.0e+00 | 0.21 (0.19-0.23) | 0.0e+00 | 0.09 (0.08-0.12) | 0.0e+00 | 0.02 (0.01-0.03) | 0.0e+00 |
| Anxiety | 0.92 (0.91-0.93) | 0.0e+00 | 0.8 (0.78-0.83) | 0.0e+00 | 0.69 (0.67-0.72) | 0.0e+00 | 0.35 (0.32-0.37) | 0.0e+00 | 1.7 (1.62-1.78) | 0.0e+00 | 3.59 (3.28-3.93) | 0.0e+00 | 7.82 (6.49-9.43) | 0.0e+00 |
| **Anxiety Male** | | | | | | | | | | | | | | |
| (Intercept) | 1.97 (1.85-2.1) | 0.0e+00 | 2.94 (2.77-3.13) | 0.0e+00 | 3.89 (3.77-4.02) | 0.0e+00 | 4.43 (4.02-4.89) | 0.0e+00 | 0.24 (0.19-0.3) | 0.0e+00 | 0.11 (0.07-0.19) | 1.1e-16 | 0.01 (0-0.03) | 1.0e-13 |
| Anxiety | 0.9 (0.87-0.94) | 4.1e-07 | 0.75 (0.69-0.81) | 6.6e-12 | 0.61 (0.53-0.7) | 6.2e-13 | 0.29 (0.25-0.33) | 0.0e+00 | 1.8 (1.54-2.09) | 4.2e-14 | 4.74 (3.71-6.05) | 0.0e+00 | 18.97 (11.14-32.31) | 0.0e+00 |
| **COVID-19-related distress** | | | | | | | | | | | | | | |
| (Intercept) | 1.92 (1.88-1.96) | 0.0e+00 | 2.81 (2.73-2.89) | 0.0e+00 | 3.88 (3.81-3.95) | 0.0e+00 | 4.35 (4.1-4.61) | 0.0e+00 | 0.22 (0.2-0.25) | 0.0e+00 | 0.11 (0.09-0.14) | 0.0e+00 | 0.02 (0.02-0.04) | 0.0e+00 |
| CV19RD | 0.97 (0.96-0.97) | 6.7e-16 | 0.93 (0.92-0.94) | 0.0e+00 | 0.93 (0.92-0.93) | 0.0e+00 | 0.72 (0.7-0.75) | 0.0e+00 | 1.32 (1.26-1.37) | 0.0e+00 | 2.03 (1.87-2.2) | 0.0e+00 | 3.23 (2.67-3.9) | 0.0e+00 |
| **COVID-19-related distress Female** | | | | | | | | | | | | | | |
| (Intercept) | 1.92 (1.87-1.96) | 0.0e+00 | 2.81 (2.72-2.9) | 0.0e+00 | 3.89 (3.81-3.97) | 0.0e+00 | 4.37 (4.08-4.68) | 0.0e+00 | 0.22 (0.19-0.24) | 0.0e+00 | 0.11 (0.09-0.13) | 0.0e+00 | 0.03 (0.02-0.04) | 0.0e+00 |
| CV19RD | 0.97 (0.96-0.98) | 5.7e-13 | 0.93 (0.92-0.94) | 0.0e+00 | 0.93 (0.92-0.93) | 0.0e+00 | 0.72 (0.7-0.75) | 0.0e+00 | 1.31 (1.26-1.37) | 0.0e+00 | 2.01 (1.84-2.2) | 0.0e+00 | 3.17 (2.58-3.9) | 0.0e+00 |
| **COVID-19-related distress Male** | | | | | | | | | | | | | | |
| (Intercept) | 1.93 (1.82-2.05) | 0.0e+00 | 2.87 (2.7-3.05) | 0.0e+00 | 3.86 (3.73-3.99) | 0.0e+00 | 4.4 (3.95-4.89) | 0.0e+00 | 0.25 (0.2-0.31) | 0.0e+00 | 0.13 (0.08-0.22) | 7.0e-15 | 0.01 (0-0.04) | 1.3e-12 |
| CV19RD | 0.96 (0.93-0.98) | 1.7e-04 | 0.93 (0.9-0.95) | 3.4e-07 | 0.92 (0.9-0.94) | 6.7e-11 | 0.72 (0.66-0.79) | 1.1e-13 | 1.32 (1.19-1.47) | 2.9e-07 | 2.1 (1.7-2.61) | 1.2e-11 | 3.59 (2.23-5.78) | 1.4e-07 |

*Table S 7 Table of Relative Risk (RR) of sleep quality and sleep quantity index using as predictors depression, anxiety, and COVID-19 related distress, adjusted for COVID-19, age, time, and recruitment type. Stratified by sex.*
